# Vδ2 T cell activation by malaria is enhanced in second infection via the cell extrinsic cytokine milieu

**DOI:** 10.64898/2026.04.29.26352021

**Authors:** Nicholas L. Dooley, Zuleima Pava, Dean Andrew, Natalie E. Stevens, Teija Frame, Jessica R. Loughland, Megan S.F. Soon, Damian A. Oyong, Fabian de Labastida Rivera, Reena Mukhiya, Julianne Hamelink, Luzia Bukali, Jessica Engel, Feargal J. Ryan, David J. Lynn, Moses R. Kamya, Isaac Ssewanyana, Rebecca Webster, James S. McCarthy, Bridget E. Barber, J. Alejandro Lopez, Chris Engwerda, Michelle J. Boyle

## Abstract

γδ T cells play critical roles in innate immunity to *Plasmodium falciparum* malaria, yet their functional heterogeneity and memory-like dynamics during first and subsequent infections remain poorly defined. Using longitudinal single-cell RNA sequencing, *ex vivo* phenotyping and *in vitro* functional analysis of γδ T cells in controlled human malaria infection (CHMI), we dissect their activation mechanisms and functions. During first infection, Vδ2 T cells dominated the responses, expanding into inflammatory and cytotoxic cells. Despite upregulation of antigen-presenting-like markers and CD16, Vδ2 T cell activation remained TCR-dependent, and these cells had no capacity to phagocytose parasites nor present antigen to CD4^+^ T cells. Leveraging a Phase I clinical study of type I IFN signalling blockade with JAK2 inhibitor ruxolitinib in CHMI, including rechallenge, we show that Vδ2 T cell activation is dependent on JAK/STAT signalling. In second malaria infection Vδ2 T cell responses are memory-like, with higher and more rapid activation, and robust induction of cytotoxic and activated terminal effector memory phenotypes. These memory-like cell responses were linked to cell-extrinsic factors, with increased systemic inflammation during second infection associated with enhanced Vδ2 T cell activation. Mechanistically we demonstrate that cytokines IL-12, IL-15 and IL-18, synergize with TCR-dependent activation by malaria parasites to enhance Vδ2 T cell responses. Memory-like Vδ2 T cells in second infection were not associated with parasite control but instead linked to inflammation and markers of disease severity. Together, these data identify key mechanisms of Vδ2 T cells activation and highlight that therapeutically targeting these cells may benefit immunopathology without compromising parasite control during malaria.

## Introduction

γδ T cells are unconventional T cells with the innate capacity to rapidly activate, producing cytokines and cytotoxic molecules upon T cell receptor (TCR) engagement. Innate activation is facilitated by the pre-programmed γδ TCR repertoire which drive responses to endogenous and pathogen-derived ligands via recognition of butyrophilin-presented phosphoantigens^1,2^, CD1-presented lipid antigens^3^, Annexin A2^4^, and other unidentified ligands. Dominating the response in humans, particularly in the peripheral blood, is the semi-invariant Vγ9Vδ2 TCR, which responds robustly to phosphoantigen-bound butyrophilin heteromers^1,2^. In addition to TCR-dependent activation, γδ T cells also respond to non-TCR signals via Toll-like-^5^, natural killer-^6^, cytokine-^7^, and inhibitory receptors^8,9^. Although γδ T cell responsiveness is limited by pre-established specificity, there is mounting evidence that these innate cells can also undergo clonal remodeling^10^ and effector memory specialization^9^, to gain memory-like functions.

In *Plasmodium falciparum* malaria^11^, Vγ9Vδ2 T cells (Vδ2 T cells) are early responders to infection, recognizing parasite-produced phosphoantigen metabolites that are released after rupture of infected red blood cells (RBCs)^12,13^. Vδ2 T cells undergo marked expansion after naturally acquired malaria in both malaria-naïve travellers and malaria-exposed individuals with uncomplicated disease ^14–18^. IFNγ and TNF production by γδ T cells has been associated with protection from high parasitemia, clinical disease and subsequent infection in some studies^19,20^. However, elevated IFNγ and TNF have also been associated with severe malaria^21–23^, and inflammatory Vδ2 T cells are increased in children with severe disease^21^ and are associated with increased risk of clinical malaria once infected^20,24^. In malaria, Vδ2 T cells also have increased expression of the cytotoxic molecules granulysin, granzyme-B and perforin^10,25–27^ and can kill intracellular and extracellular parasites in a TCR-dependent manner ^25,26,28,29^. In addition, Vδ2 T cells resemble antigen-presenting cells (APCs) in malaria, characterized by upregulation of MHC class II molecules and the co-stimulatory ligand CD86^18,27^. APC-like Vδ2 T cells have been reported to expand in response to malaria parasites *in vitro* and drive differentiation of naïve T cells^18^.

Along with these innate cell functions, memory-like adaptations in Vδ2 T cell changes are evident in children living in malaria-endemic regions, where repeated parasite exposure is associated with the modulation Vδ2 T cell inflammatory responsiveness^3030–32^, through unknown mechanisms. ^30–32^. In high-burden areas, cumulative malaria infection results in reduced Vδ2 T cell frequencies and inflammatory cytokine production in response to parasites, that is associated with asymptomatic parasite infection ^20,24^. Alongside declining inflammatory Vδ2 T numbers, frequent malaria infection is associated with the expansion of CD16^+^ (FcγRIII^+^) cells, with elevated expression of regulatory receptor TIM-3, cytotoxic molecules granulysin, granzyme B and perforin, the chemokine receptor CX3CR1 and increased inhibitory NK receptor expression^33,34^. CD16^+^ Vδ2 T cells can be activated via CD16-mediated recognition of antibody opsonized parasites, resulting in IFNγ production and parasite-directed degranulation independent of TCR engagement^34^. Non-Vδ2 γδ T cells, particularly Vδ1 T cells, also expand with cumulative malaria and exhibit clonal TCR focusing and increased proportions of effector phenotypes, which has been associated with control of symptoms in experimental infection^10^. Together, these data show that while γδ T cells have a clear role in the innate response to malaria, repeated malaria infection also drives major changes to the phenotype and function of these cells.

To date, the diverse functions of γδ T cells in malaria have mostly been studied separately, and comprehensive examination has not been undertaken of the functions induced in first exposure to malaria. Further, how rapidly γδ T cells develop memory-like responses, the mechanisms driving these changes and the consequences for protection are poorly understood. Here we leveraged control human malaria infection (CHMI) cohorts to study γδ T cells in a first infection at the transcriptional, phenotypic and functional levels. We investigate the role of type I IFN signaling in γδ T cell activation during malaria and the link between changes in the global cytokine milieu with repeated exposure, induction of memory-like Vδ2 T cells, and parasite control and pathogenesis in second infection.

## Results

### Diverse γδ T cell transcriptional profiles identified during first infection with scRNAseq

To investigate γδ T cell activation during malaria, we performed single-cell RNA sequencing (scRNA-seq) on γδ T cells isolated from volunteers (*n* = 4) enrolled in a blood-stage *P. falciparum* CHMI trial^35^ (Table S1). γδ T cells were sequenced at four timepoints: infection (I, 0 days post infection (d.p.i.)), anti-malarial drug treatment (T, 8 d.p.i.), and seven and twenty days post treatment (T+7, 15 d.p.i., T+20, 28 d.p.i.) (Fig. 1A, S1A). Following quality control and integration based on donor and timepoint, analysis of the 31,409 cells identified 14 unique clusters (Fig. 1B, S1B/C). *TRDV* and *TRGV* gene expression seen in clusters was consistent with productive recombination of known γδ T cell receptor pairs^36^ (Fig. 1C, S1D). Vγ9Vδ2 was the dominant TCR subtype (c7-c14), consistent with the abundance of this subset in peripheral blood (Fig. 1C, S1D). Less common TCR pairings, including Vδ1 paired with a range of γ chains (Vγ3, Vγ4, Vγ8, and Vγ9) and a rare Vγ8Vδ3 subset^37^ were also identified (Fig. 1C).

**Figure 1:**
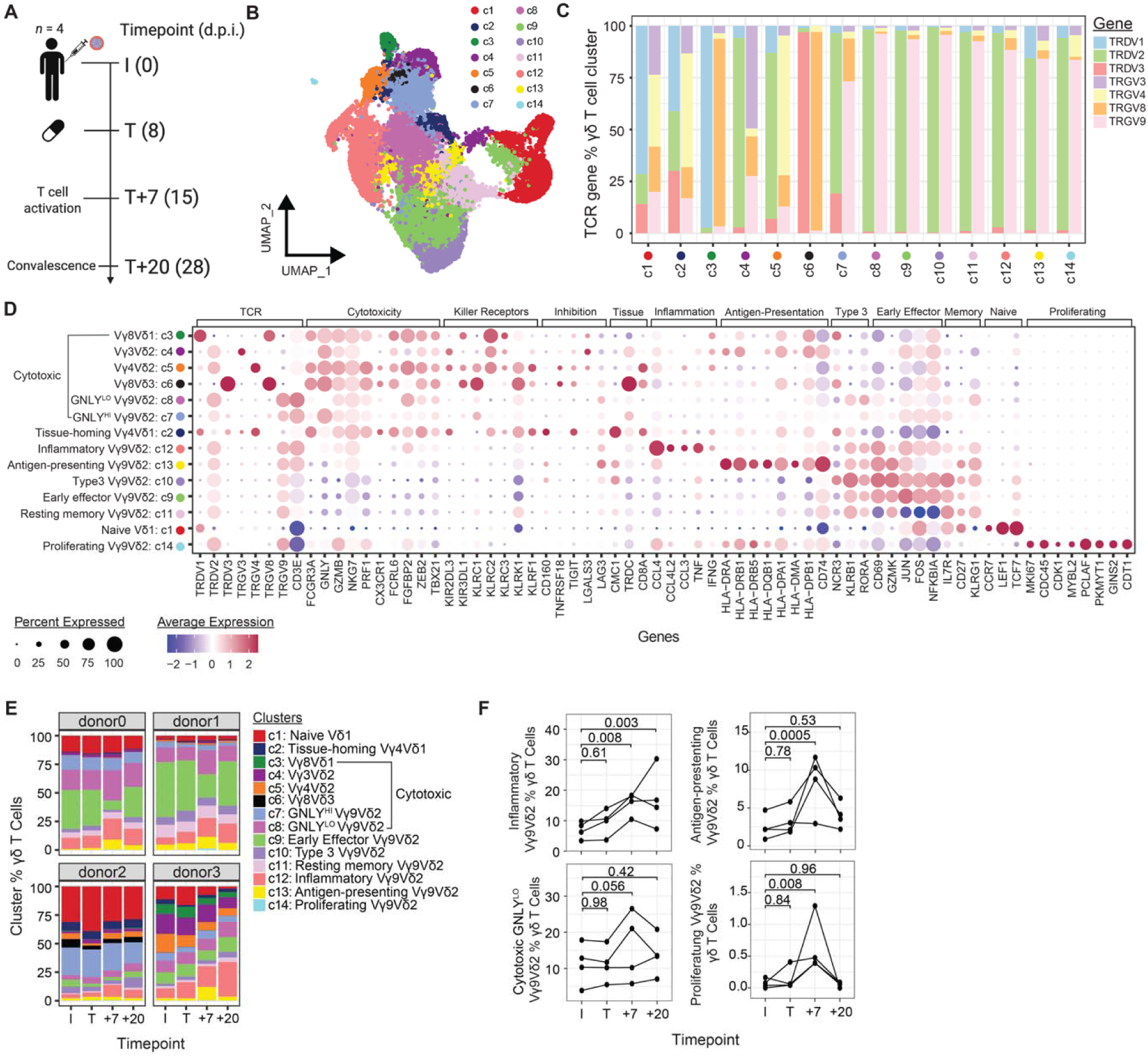
Transcriptional profiling of γδ T cells during Controlled Human Malaria Infection. (**A**) Malaria-naive volunteers (*n* = 4) were intravenously infected with *Pf*-infected RBCs in a CHMI. Venous blood drawn at infection (I, 0 days post infection (d.p.i.)), treatment (T, 8 d.p.i.), 7 (T+7, 15 d.p.i.) and 20 (T+20, 28 d.p.i.) days post-treatment. Single cell RNA sequencing libraries were constructed from isolated γδ T cells. Schematic created with BioRender.com. (**B**) UMAP of γδ T cells (31 409) identified 14 unique γδ T cell clusters. (**C**) Proportion of cells expressing TRGV and TRDV genes in each cluster. (**D**) Expression of key functional genes across annotated clusters. (**E**) Clusters as a proportion of total γδ T cell across timepoints in each donor. (**F**) Cluster proportions by donor across timepoints. P value are FDR from linear mixed effects model. See also Supplementary Fig. S1/2 and Table S2.

γδ T cell clusters were annotated based on TCR subtype and differentially expressed genes (DEGs) (Fig. 1D, Table S2). Amongst Vγ9Vδ2 clusters, two (c7 & c8) had cytotoxic phenotypes with high expression of *FCGR3A* (CD16), *GZMB* (granzyme B), *NKG7*, *PRF1* (perforin), *CX3CR1* and differing expression of *GNLY* (granulysin) (Fig. 1D, S2A/B). Granulysin has been shown to be indispensable for parasite growth inhibition and killing^25,28,29^. Cytotoxic Vγ9Vδ2 T cells expressing low levels of *GNLY* (c8; *GNLY*^LO^) had significantly higher expression of cytotoxic genes (*GZMB* and *PRF1)*, genes related to TCR activation (*CD3E*, *TRGV9* and *TRDV2*) and terminal effector genes (*KLRG1*, *ZEB2* and *TBX21*) compared to Vγ9Vδ2 T cells expressing high levels of *GNLY* (c9; *GNLY*^HI^) (Fig. 1D, S2A/B). Other Vγ9Vδ2 T cell clusters were annotated as inflammatory (c12; *CCL4*, *TNF* and *IFNG*), antigen-presenting (c13; *HLA-DR* genes and *CD74*), type 3 (c10; *NCR3*, *KLRB1* and *RORA*^38^), early effector (c9; *CD69,GZMK*) and resting memory (c11; *IL7R*, *KLRG1* and *CD27*) (Fig. 1D). A small proportion of proliferating Vγ9Vδ2 (c14) were also identified, which expressed genes involved in proliferation/differentiation, cell cycle progression and DNA replication (*MKI67*, *CDC45*, *CDK1, PCLAF, GINS2* and *CDT1*) (Fig. 1D). Amongst non-Vγ9Vδ2 clusters, four were annotated as cytotoxic (c3-c6), a Vγ4Vδ1 T cell cluster which had high expression of genes related to liver and spleen homing (c2; *CMC1*, *CD8A* and *TRDC*^39^) and another annotated as naive Vδ1 T cells (c1; *CCR7*, *CD27*, *LEF1* and *TCF7*^40^) (Fig. 1D).

The individual composition of γδ T cells was diverse; however, most clusters were present across participants and time points (except for the Vγ8Vδ3 chain recombination in c6, which was exclusive to a single participant) (Fig. 1E). Following infection, the proportion of inflammatory Vγ9Vδ2 (c12), cytotoxic *GNLY*^LO^ Vγ9Vδ2 (c8), antigen-presenting Vγ9Vδ2 (c13), and proliferating Vγ9Vδ2 cells (c14) increased significantly by T+7 (Fig. 1F, S2C). Data show high diversity of human γδ T cells with scRNAseq, however specific functional Vγ9Vδ2 T cells subtypes are preferentially activated during malaria.

### Longitudinal transcriptional activation of functionally diverse Vδ2 T cells during malaria infection

To investigate effector responses transcriptionally within γδ T cells during malaria, clusters were grouped by dominant TRDV gene expression as Vδ1, Vδ2, and Vδ3-expressing cells and malaria-induced DEGs identified for Vδ1 and Vδ2 T cells (I vs post infection timepoints) (Fig. 2A/B, Table S3). Vδ3 cells were not analyzed due to low cell counts and detection in only one participant. The largest number of DEGs were detected in Vδ2 T cells at T+7, with 145 genes upregulated and 71 genes downregulated compared to baseline (Fig. 2C). Consistent with the expansion of multiple distinct Vγ9Vδ2 cell clusters (Fig. 1F), upregulated Vδ2 genes were associated with diverse functions including proliferation (*MKI67, CDK1*), activation (*CD2*, *CD38*), inflammation (*CCL4, IFNG, TNF*), immune regulation (*CTLA4, LAG3*), cytotoxicity (*GZMB, PRF1*), antigen presentation (*HLA-DPA1*, *HLA-DPB1*) and endocytosis (*CHMP2A*, *AP2S1*) (Fig. 2D). Within Vδ1 T cells, limited DEGs were identified, but there was upregulation of *TNFSF8* (CD30-L), *CCL4* and *GZMB* at T+7 (Fig. 2D). Genes upregulated in Vδ2 T cells were expressed by cytotoxic and inflammatory γδ T cell effector clusters and highly expressed in the antigen-presenting and proliferating Vγ9Vδ2 T cell clusters at T+7 compared to I (Fig. 2E). Additionally, upregulation of a strong interferon (IFN) stimulation gene signature was detected at T+7 (*JAK3*, *JAK2*, *STAT1*, *MAP3K8*, *IFI16*, *IFI35* and *IFI27L2*) and expressed across clusters (Fig. 2D/E). Single cell pathway analysis (SPCA^41^) confirmed DEGs analysis and identified the top pathways upregulated at T+7 compared to I, including IFN signaling, interleukin signalling, TCR signaling, antigen processing and presentation and parasite infection pathways (Fig. 2F). Together, data demonstrate that Vδ2 T cells have diverse functional potential and rapidly respond during a first malaria infection.

**Figure 2:**
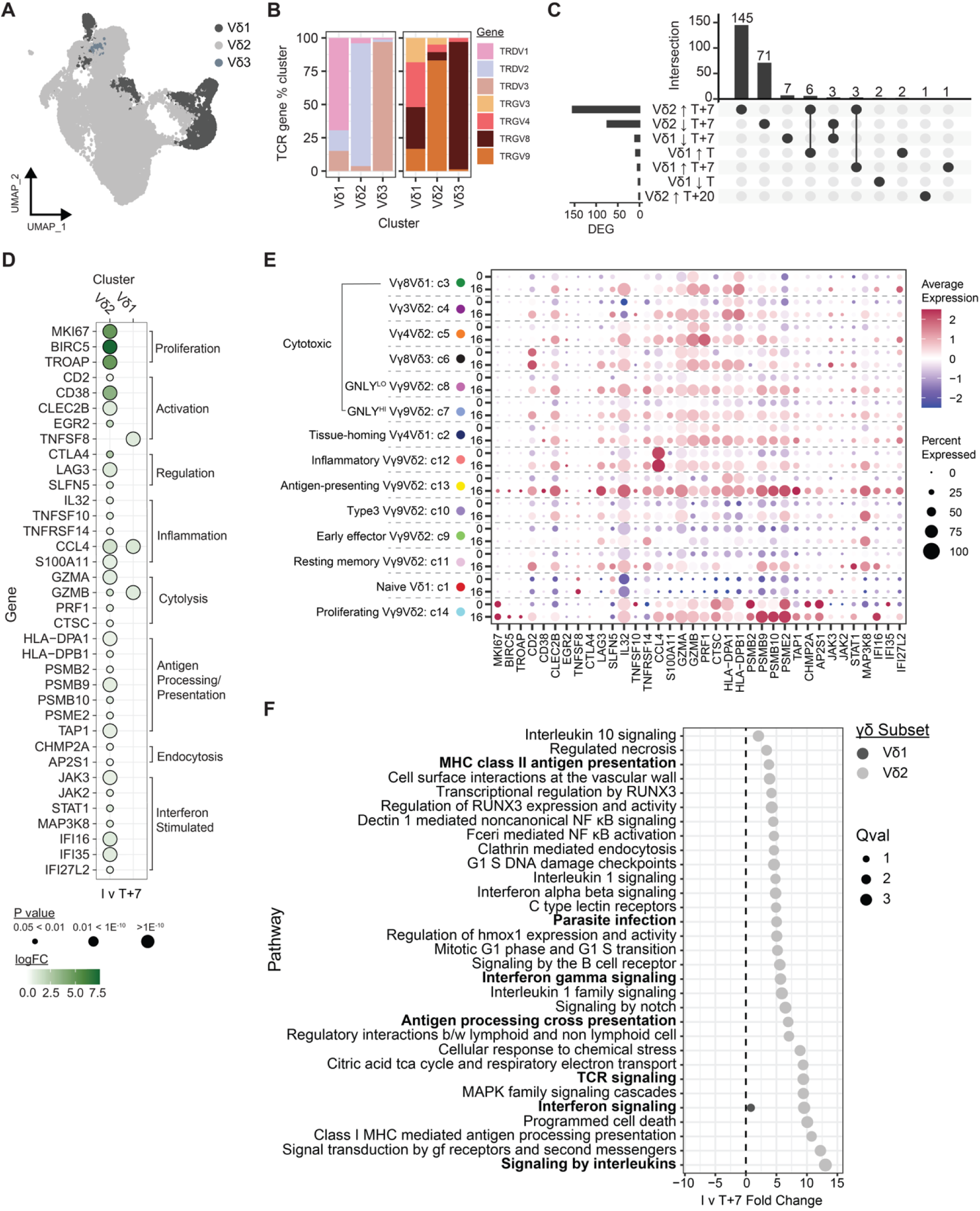
Longitudinal transcriptional changes in γδ T cells during Controlled Human Malaria Infection. (**A**) UMAP of γδ T cells analyzed with scRNAseq at I, T, T+7 and T+20 grouped by dominant TCR δ gene expression into subsets Vδ1, Vδ2 and Vδ3. (**B**) TRDV and TRGV gene proportions of TCR δ clusters. (**C**) Number of DEGs in Vδ1 and Vδ2 clusters post-infection compared to baseline (I) from pseudobulk analysis. Intersection indicates DEGs unique and shared between each cell type and timepoint. (**D**) Top DEGs T+7 compared to I in Vδ1 and Vδ2, grouped by function. (**E**) Expression of DEGs at T+7 in γδ T cell clusters at I and T+7. (**F**) Single Cell Pathway Analysis of Vδ1 and Vδ2 at T+7 compared to I. See also Supplementary Table S3.

### Vδ2 T cells expand, are activated and have increased innate cytotoxic capacity in first infection

To explore links between transcriptional activation with changes in functional capacity, we examined γδ T cell activation, phenotype and function in additional participants during CHMI^42,43^ (Fig. S3A/B, Table S1). The frequency of Vδ2 T cells increased dramatically after infection, while there was no expansion of Vδ1 T cells (Fig. 3A). Consistent with transcriptional data, robust Vδ2 T cell activation (quantified by CD38 and ICOS expression) was detected at T+7, when there was a shift in phenotypes towards effector memory (T_EM_) and central memory (T_CM_) subsets (Fig. 3B). Activation was detected in Vδ1 T cells, along with a shift towards T_CM_, despite minimal activation detected transcriptionally and no expansion of Vδ1 T cell longitudinally (Fig. 3C). Consistent with the proportional expansion and activation of inflammatory and cytotoxic Vδ2 T cells detected transcriptionally, Vδ2 T cells also increased expression of CD56, along with perforin, granzyme B, granulysin and NKG7 at T+7 (Fig. 3D, S3C/D, Table S1). Additionally, CD16, which enhances Vδ2 T cell function through antibody-mediated activation^34^, was increased (Fig. 3D). These cytotoxic changes were largely absent in Vδ1 cells (here gated as Vδ2- γδ), aside from a small increase in expression of CD16 and perforin at T+7 (Fig. 3E).

**Figure 3:**
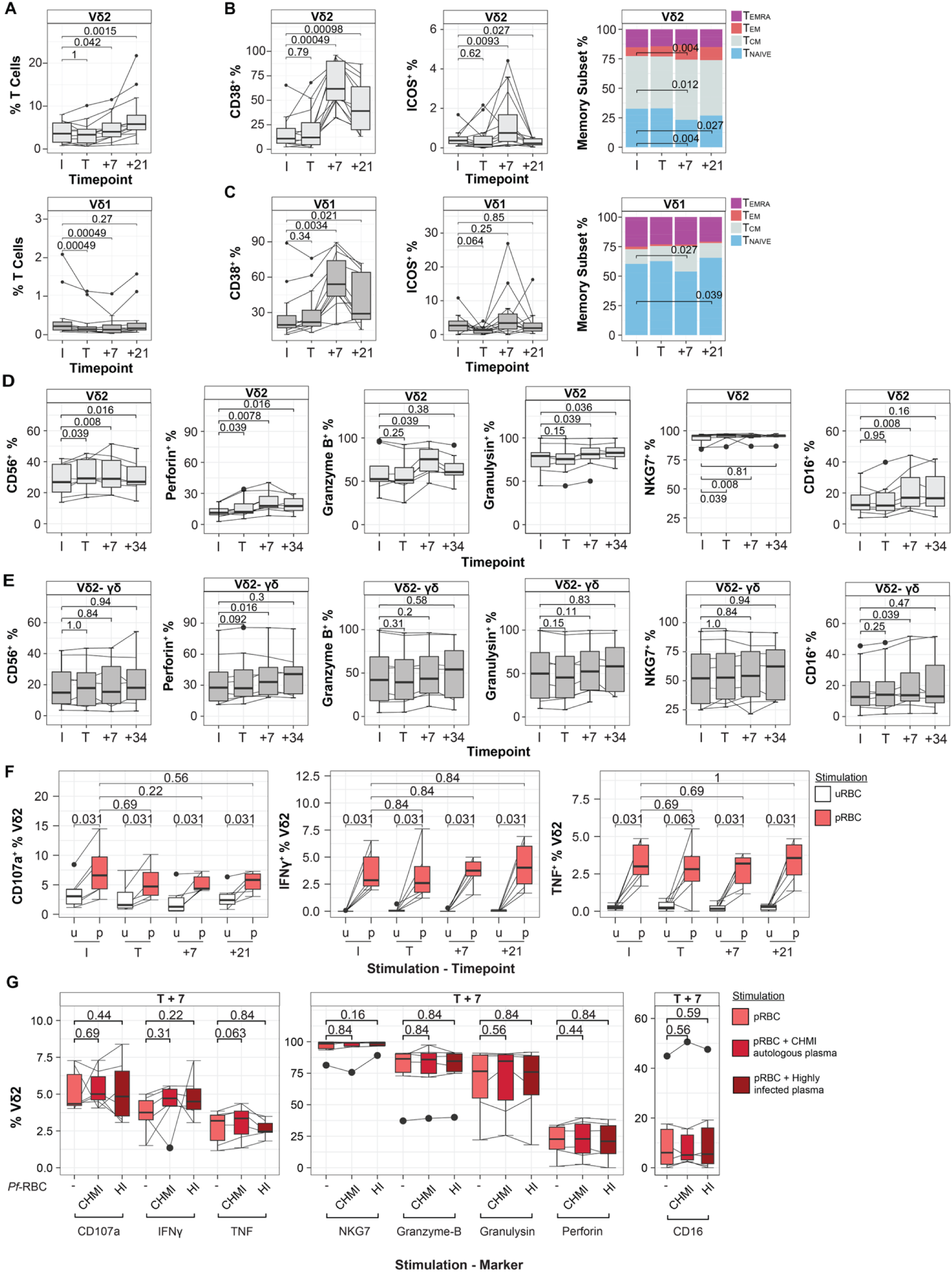
Innate cytotoxicity of γδ T cells during Controlled Human Malaria Infection. γδ T cells were analysed in PBMCs from participants during CHMI at infection (I, 0 days post infection (d.p.i.)), treatment (I, 8 d.p.i.), 7 (T+7, 15 d.p.i.) and 21/34 (T+21/34, 29/42 d.p.i.) days post-treatment. (**A**) Vδ1 and Vδ2 T cell frequency of total T cells. (**B**) Vδ2 and (**C**) Vδ1 T cell expression of CD38, ICOS and memory subset proportions (TEMRA [CD27^-^CD45RA^+^], TEM [CD27^-^CD45RA^-^], TCM [CD27^+^CD45RA^-^] and TNAIVE [CD27^+^CD45RA^+^]). (**D**) Vδ2^+^ and (**E**) Vδ2^-^ γδ T cell expression of CD56, Perforin, Granzyme-B, Granulysin, NKG7 and CD16. (**F**) Expression of CD107a, IFNγ and TNF by Vδ2 T cells after stimulation with uRBC and pRBCs. (**G**) Expression of CD107a, IFNγ, TNF, NKG7, Granzyme-B, Granulysin, Perforin, CD16 and uptake of erythrocytes by Vδ2 T cell from CHMI participants at T+7. pRBCs were un-opsonised, opsonised with autologous plasma or opsonised with plasma from a pool of highly infected donors. For all data, centre line represents the median, box limits indicate the upper and lower quartiles, whiskers extend to 1.5 times the interquartile range. Lines represent paired observations. P are from Wilcoxon signed rank test. For A-C n=12, for D-E n=8 (1 participant missing T+34) and F-G n=6. See also Supplementary Fig. 3-5.

To assess whether activation and phenotypic changes also resulted in increased Vδ2 T cell inflammatory and cytotoxic functions in malaria, we stimulated CHMI^42^ participant cells with intact *P. falciparum*-infected RBCs (pRBCs) across the course of infection and analyzed IFN-γ and TNF accumulation, along with degranulation quantified by CD107a as a marker of cytotoxicity (Fig. S4A/B, Table S1). Vδ2 T cells had robust innate effector responses at all timepoints, with increased CD107a expression and production of IFNγ and TNF upon stimulation with intact pRBCs compared to uninfected RBCs (uRBCs) (Fig. 3F). No functional responses were detected from Vδ2- γδ T cells (Fig. S4C). The magnitude of the Vδ2 T cell response did not change during or after *P. falciparum* infection, with no changes in proportions of degranulation, cytotoxic molecule expression, nor cytokine production (Fig. 3F, S4D). In previous studies, Vδ2 T cells from malaria-naïve donors have reported to not degranulate in response to parasites in the absence of *in vitro* cytokine priming^26^. However, our findings indicate robust activation of Vδ2 T cells within PBMCs when incubated with parasites regardless of malaria exposure, indicative of an innate cytotoxic response.

### Antibody dependent cellular cytotoxicity is not acquired by Vδ2 T cells during first infection

Vδ2 T cells can acquire antibody-dependent cell cytotoxicity (ADCC), inflammatory and phagocytic functions mediated via CD16 engagement, either following repeated infection or after *in vitro* expansion with IL-2 and IL-15^26,34^. Because CD16 expression was upregulated in Vδ2 T cells during CHMI (Fig. 3D), we investigated if this resulted in enhanced antibody dependent functions to pRBCs opsonized with plasma from CHMI participants at matched time points or from a pool of highly exposed Ugandan individuals^44^. Compared to un-opsonized pRBC, opsonization with autologous plasma from CHMI participants or from Ugandan individuals did not enhance Vδ2 T cell degranulation, inflammatory cytokine production, or accumulation of cytolytic proteins and proteases before, during or following infection (Fig. 3G, S5). Additionally, CD16 expression remained unchanged following incubation with opsonized parasites (Fig. 3G). Together, these findings suggest that Vδ2 T cell degranulation occurs independently of Fc receptor signaling, and indicate that Vδ2 T cells exert innate, antibody-independent cytotoxic responses against blood-stage *P. falciparum* in first infection.

### APC-like Vδ2 T cells induced by malaria do not have functional capacity

APC-like functions of Vδ2 T cells have been reported in malaria, where following culture with parasites, Vδ2 T cells expressed HLA-DR, CD80, CD83, CD86, and CD40 and can drive CD4 and CD8 T cell differentiation^45^. Similar APC-like functions of γδ T cells have been described in response to *Escherichia coli*^46^, *Mycobacterium tuberculosis*^47^ and *Listeria monocytogenes*^48^. Consistent with transcriptional analysis, we confirmed increased expression of HLA-DR and the co-stimulatory receptor CD86 on Vδ2 T cells at T+7 (Fig 4A, S6). Further, culturing PBMCs with pRBC lysate or the Vδ2 TCR agonist HMBPP induced HLA-DR, CD86 and CD40 expression on Vδ2 T cells compared to controls (media alone or stimulated with uRBC lysate) or *ex vivo* (Fig. 4B, S7). However, we found that parasite cultured γδ T cells did not drive CD4^+^ T cell proliferation beyond controls (Fig. 4C, S8). Further, we observed no internalization of un-opsonized or opsonized intact pRBC by Vδ2 T cells at any timepoint during infection, even though myeloid cells in the same assay readily internalized intact pRBC opsonized with plasma from highly exposed Ugandan individuals (Fig. 4D, S9). Thus, our results show little evidence of APC-like γδ T cell function in malaria, despite upregulation of genes and proteins associated with APCs.

**Figure 4:**
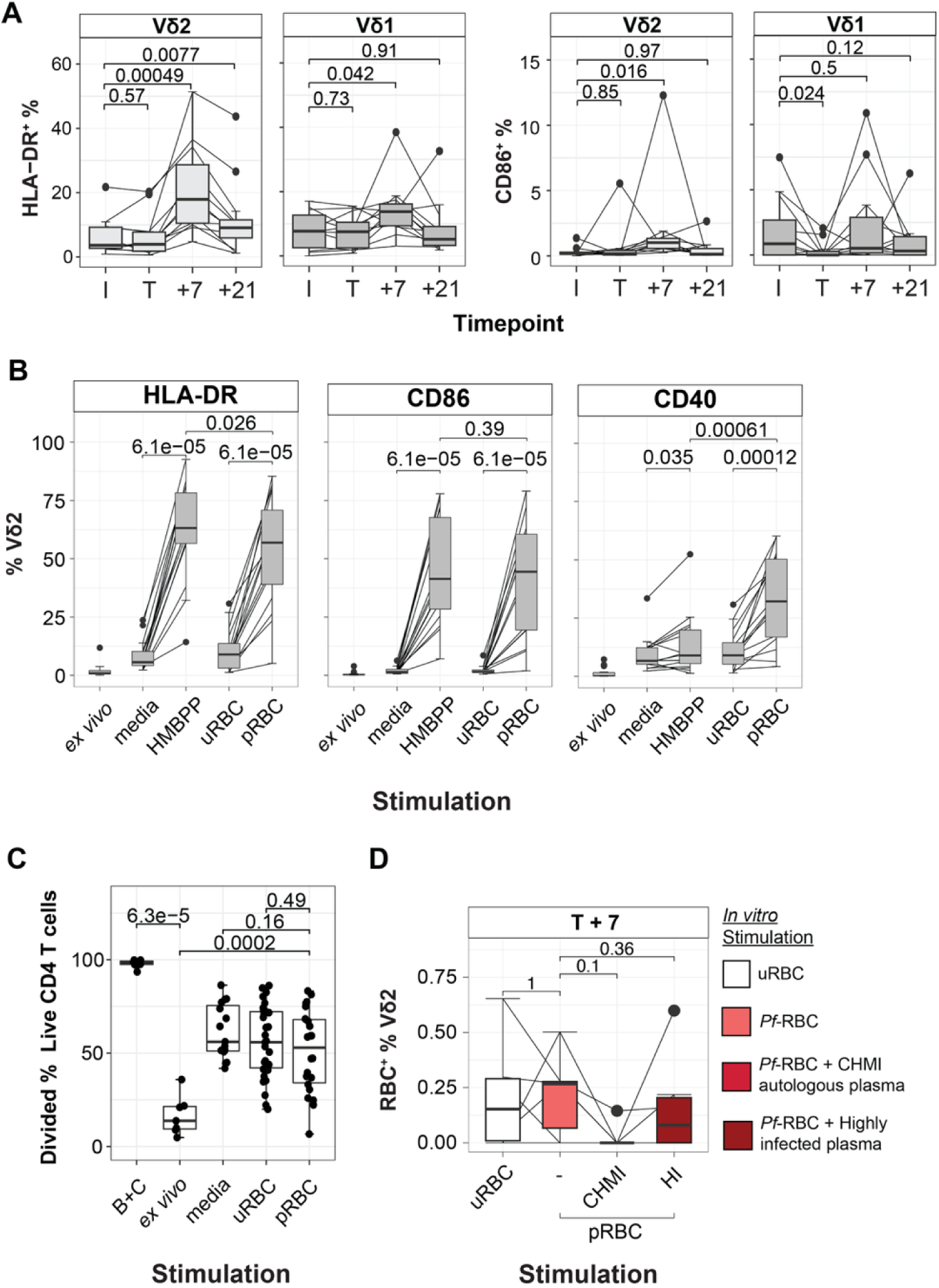
Antigen-presenting capacity of γδ T cells during Controlled Human Malaria Infection. **A**) Vδ2 and Vδ1 T cell expression of HLA-DR and CD86 during CHMI in PBMCs at infection (I, 0 days post infection (d.p.i.)), treatment (T, 8 d.p.i.), 7 (T+7, 15 d.p.i.) and 21 (T+21, 29 d.p.i.) days post-treatment (*n* = 12). (**B**) HLA-DR, CD86 and CD40 expression on Vδ2 T cells from malaria-naïve donors (*n* = 15) ex vivo and after culture for 5 days with media alone, HMBPP, uRBC lysate and pRBC lysate. (**C**) Activation of isolated naïve CD4 T cells with γδ T cells (*n* = 6) isolated *ex vivo*, or after 3-day culture with media, uRBC lysate or pRBC lysate. Positive control is CD4 T cells with CD3/CD28 beads and IL-12, IL-23 and TGF-β (B+C). Frequency of divided cellS in CD4 T cell population after co-culture is shown. (**D**) Phagocytosis of uRBC or pRBC stained with Cell Trace Violet in Vδ2 T cells (n=6) from PBMCs at T+7 during CHMI. Centre lines represent median, box limits indicate the upper and lower quartiles, whiskers extend to 1.5 times the interquartile range. Lines represent paired observations. P from paired data are Wilcoxon signed rank tests. Unpaired comparisons made with Mann-Whitney U test. See also Supplementary Figures 6-9.

### Vδ2 T cell activation in malaria is JAK/STAT-dependent

Having analyzed functions of Vδ2 T cells induced during a first malaria infection, we next sought to identify pathways and mechanisms driving Vδ2 T cell activation. Transcriptional analysis of Vδ2 T cells during CHMI identified that activation occurred concurrently with the upregulation of IFN signaling genes. To test if IFN signaling had a direct role in Vδ2 T cell activation, we analyzed cells during CHMI where ruxolitinib, a JAK1/2 inhibitor that can prevent type I IFN signaling, was tested as a host-directed therapy ^49^. In this study, 20 participants were enrolled in a blood-stage *P. falciparum* CHMI and treated with the anti-malarial drug artemether-lumefantrine at day 8/9 post-infection, together with either ruxolitinib (*n* = 11) or placebo (*n* = 9) for three days (Fig. 5A, Table S1). Here, we used PBMC samples collected at infection (I, 0 d.p.i.), treatment (T, 8/9 d.p.i.), and 3, 7, and 21 days following treatment (T+3, T+7, T+21). Vδ2 T cell memory and effector subsets, as well as those expressing activation and functional phenotypes were quantified (Fig. S10). Data were analyzed using linear mixed effects models, comparing I to T, and then T to post-treatment with treatment group included as an interaction term to identify ruxolitinib-mediated changes, as previously ^49^.

**Figure 5:**
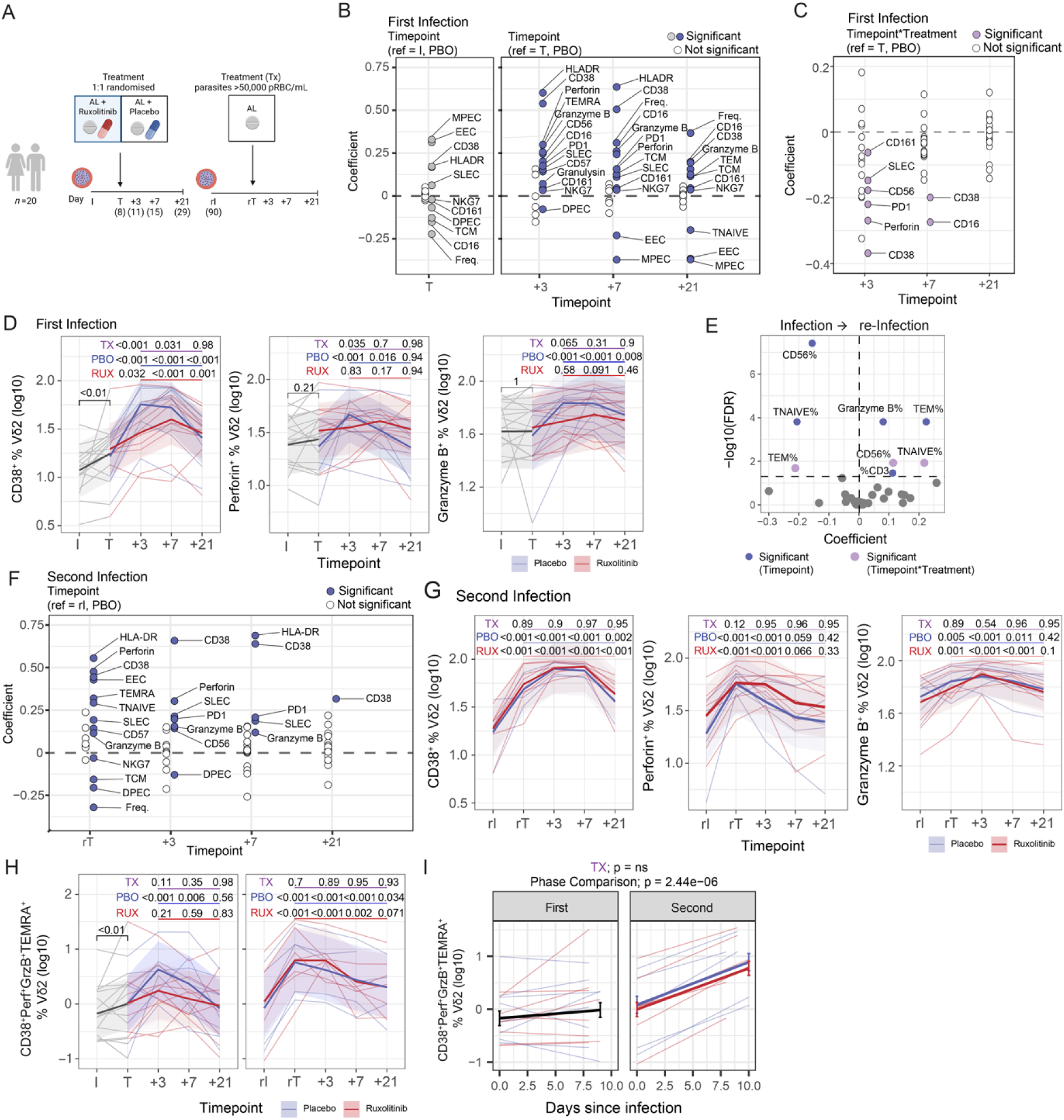
Role of Type I IFN signalling in Vδ2 T cells activation and responses in second infection. (**A**) Vδ2 T cells were analysed in volunteers (n=20) during CHMI to test ruxolitinib as a host-directed therapy. Individuals were infected (I), and at treatment with artemether-lumefantrine and ruxolitinib or placebo at (T, 8/9 d.p.i.), and then followed (T+3, T+7, T+21). At day 91 individuals were reinfected and cells analysed (rI, rT, rT+3, rT+7, rT+21). **(B–C)** Vδ2 T cell marker expression was analysed after treatment during the first infection (I vs T; T vs T+3, T+7, T+21). Linear mixed-effects model coefficients for time (in placebo-treated participants) (**B**) and for treatment interaction (impact of ruxolitinib, TX) (**C**). Blue/grey dots indicate significant coefficients for time in placebo (PBO). Purple dots indicate significant coefficients for treatment and white indicates non-significant changes. (**D**) CD38, perforin, and granzyme B expression by Vδ2 T cells during the first infection. P values are from linear mixed-effects models in PBO, ruxolitinib-treated (RUX, identified with contrasts), and for the interaction term (TX) groups. (**E**) Linear mixed-effects model coefficients for Vδ2 T cell marker expression from first to second infection (I vs rI), significant in PBO (blue) and treatment TX (purple). (**F**) Linear mixed-effects model coefficients for marker expression after treatment during the second infection (rI vs rT, rT+3, rT+7, rT+21) in placebo-treated participants. Blue indicates significant increases in PBO, purple indicates treatment TX, and white indicates non-significant changes. (**G**) CD38, perforin, and granzyme B expression during the second infection, analysed in PBO, RUX, and TX groups. (**H**) Frequency of CD38^+^perforin^+^granzyme B^+^T_EMRA_^+^ Vδ2 T cells during the first and second infection, P values are from linear mixed-effects models in PBO, ruxolitinib-treated (RUX, identified with contrasts), and for the interaction term (TX) groups. (**I**) CD38^+^perforin^+^granzyme B^+^T_EMRA_^+^ Vδ2 T cell frequency from I to T across first and second infections. P is from segmented linear regression model. See also Supplementary Figures 10–12.

Confirming the rapid activation of Vδ2 T cells during first infection, increased frequencies of CD38^+^ and HLA-DR^+^ cells were detected at day of treatment (Fig. 5B/D, S11A). Activation increased further in the placebo group at T+3, with increased frequencies of Vδ2 T cells expressing CD38, HLA-DR, CD16, perforin, granzyme B, granulysin, NKG7, CD161, CD57 and PD1 compared with day of treatment (Fig. 5B/D, S11A). The proportion of terminally differentiated memory (T_EMRA_) and short-lived effector cells (SLEC) also expanded (Fig. 5B, S11A). The elevation of many activation and cytotoxic markers remained at T+7 and T+21, and increased frequencies of Vδ2 T cells within the T cell compartment were detected at T+21 (Fig. 5B, S11A). Ruxolitinib treatment had a significant impact on Vδ2 T cell activation, with reduced induction of CD38, Perforin, PD1, CD56 and CD161 at T+3 in ruxolitinib treated individuals compared to placebo, indicating JAK-STAT signaling has a major role in Vδ2 T cell activation (Fig. 5C/D, S11A). Reduced induction of CD38 and CD16 were also detected at T+7 in ruxolitinib treated individuals (Fig. 5C/D, S11A). Together, these data reinforce the dominant role of Vδ2 T cells in first infection and show that signalling through JAK-STAT drives Vδ2 T cell activation in malaria.

### Vδ2 T cell activation is accelerated in second malaria infection

In this CHMI study, 15 individuals (ruxolitinib *n* = 9, placebo *n* = 6) were reinfected three months after the first infection and again treated with artemether-lumefantrine at a parasitemia threshold of ≥ 50,000 pRBCs/mL or when symptoms developed (day 8-10 post re-infection)^49^. Ruxolitinib was not re-administered in the second infection. To investigate if Vδ2 T cells showed memory-like responses during a second *P. falciparum* infection, we measured cellular responses during second infection at timepoints matching primary infection, re-infection (rI), re-treatment (rT), and day 3, 7 and 21 post treatment (rT+3, rT+7, rT+21) (Fig. 5A). Consistent with previous studies showing expansion of Vδ2 T cells following naturally-acquired malaria^14–18^, at re-infection, the frequency of Vδ2 T cells, along with Granzyme-B expression and the proportion of T_EM_, were higher compared to first infection (Fig. 5E).

After second infection, we detected the rapid activation of cytotoxic Vδ2 T cells through increased frequencies of CD38, HLA-DR, Perforin, Granzyme B, CD57 and PD1 expressing cells, as well as expansion of T_EMRA_ and SLEC memory subsets by rT (Fig. 5F/G, S11A), in contrast to first infection where most activation occurred 3 days after drug treatment (T+3). Vδ2 T cells co-expressed multiple activation and cytotoxic markers and assumed effector cell phenotypes by rT. Notably, CD38^+^perforin^+^granzyme B^+^T_EMRA_^+^ Vδ2 T cells rapidly expanded after second infection, making up approximately 10% of the Vδ2 T cell compartment by rT, but were far less abundant in first infection and only expanded at T+3 in the absence of ruxolitinib treatment (Fig. 5H). To formally test whether Vδ2 T cell activation and cytotoxicity increased more rapidly in second infection, we used a segmented linear regression model to compare the magnitude of activation between I and T to that between rI and rT, as previously^49^. This approach confirmed that Vδ2 T cell activation and induction of cytotoxic effector function was significantly faster in the second infection (Fig. 5I and S12). In all cases, there was no evidence that ruxolitinib treatment in first infection impacted these rapid Vδ2 T cell memory-like responses in second infection, and considering all responses over the course of second infection the only significant treatment effect detected was a minor increase in T_EM_ cells at rT+7 in ruxolitinib treated individuals (Fig. S11B). This contrasts with responses in CD4^+^ T cells, where treatment with ruxolitinib in first infection imprinted multiple differences that were observed in second infection^50^. Together, these data show that Vδ2 T cells have a marked memory-like response to a second *P. falciparum* infection, with faster activation and expression of cytotoxic molecules; this was not modulated by treatment with ruxolitinib in first infection.

### IL-12, IL-15 and IL-18 enhance Vδ2 T cell parasite-induced TCR-dependent activation

Next, we investigated the cell intrinsic and cell extrinsic mechanisms driving the rapid activation and expression of cytotoxic molecules by Vδ2 T cells in second infection. In previous analysis of circulating cytokines in this host-directed study, data indicated that the global inflammatory response was also more rapid in the second infection^49^. This rapid response included increased levels of monocyte-derived chemo-attractants CXCL9 and CXCL10, which have been associated with Vδ2 T cell expansion during malaria in other studies^16,49^. Further, circulating levels of IL-12 (IL-12p70 and IL-12B), IL-15RA and IL-18BP all increased^49^, suggesting changes to these cytokine signalling pathways in second infection; IL-12, IL-15, IL-18 are strong activators of innate lymphocytes, particularly NK cells, but have also been reported to drive Vδ2 T cell activation independently of TCR-engagement^51,52^. To assess if the more rapid Vδ2 T cell expansion could be driven by increased inflammatory milieu in the second infection, we investigated if activated cytotoxic (CD38^+^perforin^+^granzyme B^+^) T_EMRA_ Vδ2 T cells correlated with CXCL9/10, and IL-12/15/18 in the circulation. Activated cytotoxic Vδ2 T cell frequencies positively correlated with CXCL9, CXCL10, IL-12B and IL-15, but not IL-18 (Fig. 6A).

**Figure 6:**
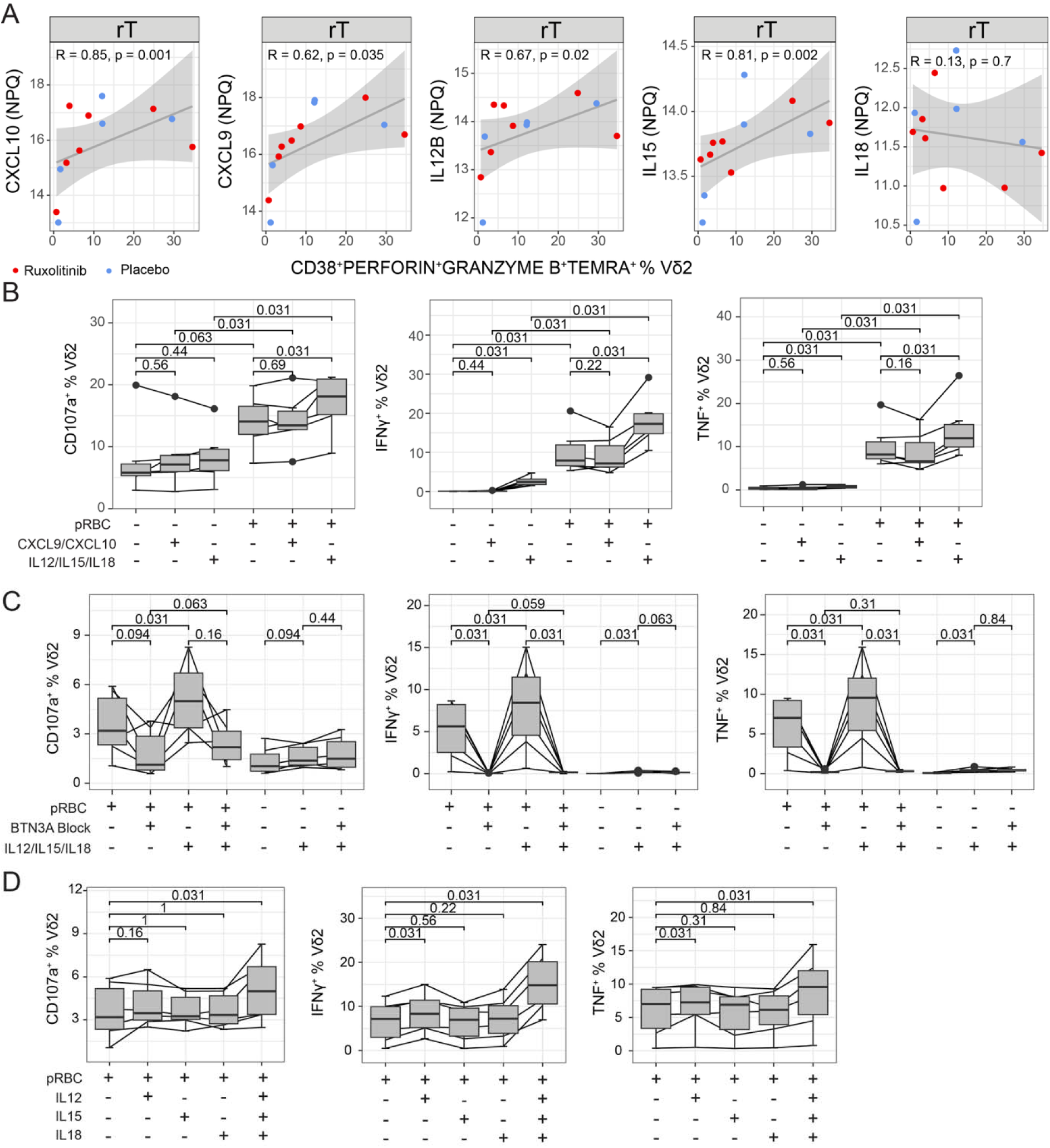
Cytokines synergise with TCR-dependent activation of Vδ2 T cells in malaria. (**A**) Association of CD38^+^perforin^+^granzyme B^+^T_EMRA_^+^ Vδ2 T cells with cytokines (Nulisa Protein Quantification) at re-treatment (rT) in second CHMI. Spearman’s Rho and p value shown. **B**) CD107a, IFNγ and TNF expression by Vδ2 T cells from malaria naïve donors (n=6) after co-culture with parasite lysate or media with and without CXCL9/10 or IL-12/15/18. (**C**) CD107a, IFNγ and TNF expression by Vδ2 T cells after co-culture with parasite lysate or media with and without IL-12/15/18 and anti-BTN3A. (**D**) CD107a, IFNγ and TNF expression by Vδ2 T cells after co-culture with parasite lysate with and without IL-12, IL-15, IL-18 and IL-12/15/18. For B-D Centre lines represent median, box limits indicate the upper and lower quartiles, whiskers extend to 1.5 times the interquartile range. Lines represent paired observations. P are from Wilcoxon signed rank test. See also Supplementary Fig. 13.

While TCR independent, cytokine mediated activation of Vδ2 T cells has been reported for combinations of IL-12, IL-15 and IL-18^51,52^, it is unknown if these cytokines can also enhance TCR dependent activation. To formally test if cytokines could contribute to the rapid activation of Vδ2 T cells in second infection by enhancing malaria driven TCR-dependent activation, we incubated PBMCs from malaria-naïve donors with pRBC lysate and/or CXCL9/10, or IL-12/15/18. Following incubation, activation of Vδ2 T cells was quantified by IFN-γ and TNF production and CD107a expression to measure degranulation (Fig S13A/B). In the absence of parasites, IL-12/15/18 alone, but not CXCL9/10, induced IFN-γ and a small amount of TNF but not CD107a in Vδ2 T cells (Fig. 6B). In combination with pRBC lysate, IL-12/15/18 enhanced IFN-γ, TNF and CD107a expression compared to pRBC lysate alone or cytokines alone (Fig. 6B). CD38 expression was also increased, and there was a small but significant decrease in granulysin expression, consistent with increased degranulation (Fig S13C). CXCL9/10 incubated with pRBC lysate did not increase activation above that seen with parasites alone (Fig. 6B).

To assess the TCR dependence of cytokine enhanced malaria activation, we quantified responses in cells incubated with pRBC lysate, IL-12/IL-15/IL-18 and α-BTN3A blocking antibody (Fig. S13D/E). Incubation with an α-BTN3A blocking antibody reduced degranulation and completely suppressed inflammatory cytokine production by Vδ2 T cells stimulated with parasite lysate alone or in combination with IL-12/IL-15/IL-18, indicating cytokines could enhance malaria TCR-dependent activation (Fig. 6C). In contrast, α-BTN3A blocking antibody did not impact degranulation and inflammatory cytokine production by Vδ2 T cells incubated with IL-12/IL-15/IL-18 alone, indicating TCR-independent activation by cytokines in the absence of parasites (Fig. 6C). To identify which of the cytokines had the greatest impact on parasite-mediated activation, each single cytokine was incubated with PMBCs. No single cytokine enhanced Vδ2 T cell degranulation, and only IL-12 increased IFN-γ and TNF production above that observed with parasite alone, suggesting synergistic activity of cytokines (Fig. 6D). Together, these data highlight a newly identified mechanism of Vδ2 T cell activation during malaria whereby the circulating cytokine milieu synergises with TCR-dependent activation to enhance function.

### Malaria does not drive epigenetic changes to Vδ2 T cells

While the above data suggest that enhanced Vδ2 T cell activation in second infection is due to increased inflammatory milieu, we also considered cell intrinsic factors driving memory-like responses by assessing chromatin accessibility. Vδ2 T cells were sort-purified from participants during CHMI at infection (I, 0 d.p.i.), treatment (T, 8 d.p.i.), and days 7, and 19 following treatment (T+7, T+19) and analyzed by ATAC-seq (*n* = 6, Fig. S14A and Table S13). Initial PCA analysis of the peak counts indicated participant and/or batch differences, which were controlled for in subsequent analysis. We found little evidence of malaria-driven chromatin changes, and Vδ2 T cells did not cluster by day after controlling for participant and batch variation (Fig. S14B). Indeed, differential accessibility analysis comparing day 0 to subsequent timepoints did not identify any regions that were differentially accessible after correcting for multiple comparisons (Table S14). Further, even if considering regions with a modest change (0.25-fold) and FDR of 0.15 or less, no regions with clear roles in activation were identified (Table S14, Figure S14C). Thus, there was little evidence that malaria drives epigenetic changes to Vδ2 T cells that can explain enhanced activation in second infection, consistent with cell extrinsic factors underpinning memory-like responses.

### Memory-like Vδ2 T cell response is not associated parasite control, but instead drives inflammation

We previously reported evidence of immune acquisition resulting in lower parasite multiplication rates (PMR_48hr_) in both placebo and ruxolitinib treatment groups in second infection in this CHMI^49^. To assess if the memory-like Vδ2 T cells in second infection contributed to parasite control, we investigated correlations with PMR_48hr_ and peak parasitemia. However, there was no correlation between PMR_48hr_ or parasitemia and frequencies of CD38^+^perforin^+^granzyme B^+^T_EMRA_^+^ Vδ2 T cells (Fig. 7A/B), suggesting these cells did not have a role in controlling parasites.

**Figure 7:**
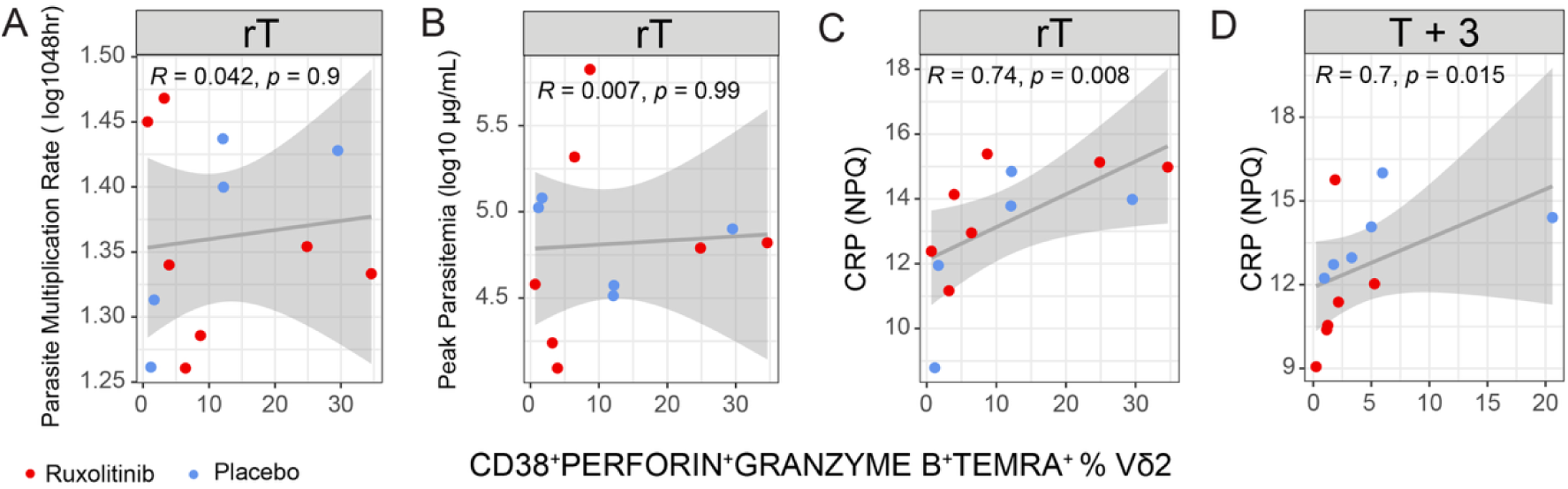
Association between with activated cytotoxic Vδ2 T cells and parasitemia or CRP. Correlation between CD38^+^perforin^+^granzyme B^+^T_EMRA_^+^ Vδ2 T cell frequency with (**A**) parasite multiplication rate, (**B**) peak parasitemia, (**C/D**) C-reactive protein (CRP, Nulisa Protein Quantification) at indicated timepoints. Spearman’s Rho and p value shown. See also Supplementary Fig. 15.

In contrast to reduced parasite multiplication, in second infection multiple markers of inflammation were increased^49^. Because inflammatory responses from Vδ2 T cells are associated with severe malaria^21,24^, we also considered a role of memory-like Vδ2 T cells in inflammation by examining associations with the key marker of disease severity, C-reactive protein (CRP). CRP increases more rapidly in second infection^49^, peaking by time of treatment (rT) in many individuals, in contrast to first infection where the peak response occurs at T+3 (in the placebo group) (Fig S15). In second infection, CRP was strongly associated with CD38^+^perforin^+^granzyme B^+^ T_EMRA_^+^ Vδ2 T cell frequency at rT, suggesting a role of Vδ2 T cell activation in disease (Fig. 7C). CRP and the frequencies CD38^+^perforin^+^granzyme B^+^T_EMRA_^+^ Vδ2 T cells were also correlated at T+3 in first infection (Fig. 7D), and both CRP and activation of Vδ2 T cell were reduced by ruxolitinib treatment at this time point (Fig. S15, Fig. 5C). Together data highlight that despite gaining memory-like responses, Vδ2 T cells remain drivers of pathogenesis in second infection

## Discussion

γδ T cells are a key ‘semi-innate’ lymphocyte population, with multiple roles in infection and disease. During malaria, Vδ2 T cells are major inflammatory responders which may have roles in protection from infection^20^, but have also been associated with severity of disease in children^20,21,24^. After repeated exposure to *P. falciparum*, Vδ2 T cells show reduced proliferation and inflammatory cytokine production, but increased CD16 expression which mediates antibody-dependent activation. Here, data from CHMI show that in a first malaria infection, Vδ2 T cell activation occurs via type I IFN signaling, which drives expansion of a specific subsets of Vδ2 T cells which produce IFNγ and TNF, but also have cytotoxic functions, and degranulate in response to parasites. In second infection, activation of Vδ2 T cells is more rapid, mediated by a previously unrecognised mechanism whereby the myeloid-derived cytokine milieu, particularly IL-12/IL-15/IL-18, synergize with TCR-dependent pathways to enhanced activation. This memory-like Vδ2 T cell response was not associated with parasite control but instead strongly correlated with markers of disease. Together, these data identify key mechanisms of Vδ2 T cells activation and highlight that therapeutically targeting these cells may benefit immunopathology without compromising parasite control.

Vδ2 T cells are primarily activated by TCR recognition of conformational changes in BTN3A1/2A1 on host cells^1,2^, which in malaria are triggered by release of phosphoantigen metabolites secreted by ruptured parasite-infected RBCs^12,13^. Here we show that this TCR dependent response is also modulated by the global environmental. We have previously reported that the global cytokine milieu is marked by a significantly more rapid inflammatory response in second malaria infection ^49^. Many of these rapidly induced responses were myeloid derived, and here we show that IL-12/IL-15 and IL-18, typically associated with NK cell activation, drive the emergence of a memory-like activation profile of Vδ2 T cells by synergising with TCR-dependent activation. In contrast to this enhanced response in second infection, in areas of chronic malaria exposure Vδ2 T cells have reduced proliferation and inflammatory responses to parasites, presumably due to reduced TCR-dependent activation. These changes have been linked to remodelling of the Vδ2 T cell compartment, including increased expression of regulatory receptors such as Tim-3, CD57 and regulatory KIRs^6,9,20,24,34^. However, our data suggest that global changes to other immune cells should also be considered. Indeed, parasite induced myeloid derived cytokines IL-12/IL-15 and IL-18 are likely modulated by repeated exposure. Children in areas of high malaria transmission have expansion of non-classical monocytes^53^, which we have shown do not produce cytokines in response to parasites^54^. Further, monocytes become attenuated with age in malaria endemic areas^55^, and malaria drives epigenetic reprogramming of innate cells resulting in modulated cytokine responsiveness ^56^. This reduced response from monocytes may thus limited synergising impact of cytokines on TCR-dependent activation of Vδ2 T cells. This mechanisms of cytokine synergising with TCR-dependent Vδ2 T cells activation and modified myeloid populations may also underpin memory-like Vδ2 T cells in other diseases. For example, memory Vδ2 T cells have been reported following BCG vaccination^57^. However, BCG is a well-recognized driver of innate cell memory, or trained immunity, which enhances the inflammatory response of monocytes ^58^. Thus, our data highlight that the global cytokine milieu should be considered in other settings where Vδ2 T cell ‘memory’ responses have been observed. Similarly, changes to the global cytokine milieu have also been linked to modulated NK cell activation in malaria^59^, highlighting the intricate interplay between cell types in the host immune response to infection.

Importantly, despite the memory like response of Vδ2 T cells in second infection, there was no evidence that these cells had a role in controlling parasite growth, which we previously reported was lower in second infection ^49^. This finding may be due to the specific phenotypes of Vδ2 T cells induced, which do not increase granulysin, nor gain antibody-dependent functions. Granulysin is required for TCR-dependent parasite killing by Vδ2 T cells^25,28,29^. Granulysin expression was not increased in Vδ2 T cells second infection, nor did the granulysin-high (*GLNY*^HI^) transcriptional cluster expand in first infection. Further, antibody-dependent activation and degranulation^34^, and CD16-mediated opsonic phagocytosis has been proposed to control parasites^26^, and is increased in children from high transmission areas. However, despite identifying APC-like Vδ2 T cell phenotypes, with increased CD16 expression during CHMI, Vδ2 T cells did not have FcγR-dependent responses, nor the ability to phagocytosis parasites. Thus, the major remodeling of Vδ2 T cells to gain Fc-dependent functions reported in children in high malaria endemic areas does not appear to occur after one or two infections. Instead, Vδ2 T cells have potent innate inflammatory and cytotoxic effector activity in both first and second infection. This response was associated with CRP, a key marker of inflammation, suggesting a major role of Vδ2 T cells in pathogenesis. These findings are consistent with studies in natural malaria infection where inflammatory Vδ2 T cells are associated with increased disease severity^20,21^, as well as recent findings that report malaria activated γδ T cells contribute to blood-brain-barrier (BBB) breakdown in in vitro human models of cerebral malaria^60^. Importantly, we show that Vδ2 T cell activation is dependent on type I IFN and JAK/STAT signaling, and ruxolitinib treatment can reduce Vδ2 T cell activation and associated inflammation. Ruxolitinib has also been shown to prevent parasite-driven pathogenesis in 3D-BBB models^60,61^, thus this host-directed therapy is an exciting avenue to improve severe disease by directly preserving vascular integrity as well as by reducing immunopathology.

Limitations which should be considered in interpreting our findings include the mild and short duration infections in these CHMI studies where early treatment of participants is required for safety. Additionally, these studies are restricted to adults, and age-related differences in Vδ2 T cells in children may be an important consideration in naturally acquired malaria^54^. Within our scRNAseq analysis we did not investigate γδ T cell TCRs with V(D)J sequencing; however, a previous study has shown limited change in clonal diversity among participants across four CHMI trials^10^. Our study also investigated γδ T cells responses in the peripheral blood and while this is relevant to blood-dominant Vγ9Vδ2 T cell, further studies of tissue where other γδ T cells dominate are required.

## Methods

### Ethics

Written informed consent was obtained from all participants. Ethics approval for the use of human samples was obtained from the Alfred Human Research and Ethics Committee for the Burnet Institute (#288/23, #166/24, #328/17), the Human Research and Ethics Committee of QIMR Berghofer (P1479 and P3696, ), the Makreree University School of Medicine Reseach and Ethics Committee (2011-167), the Ugandan National Council of Science and Technology (HS1019), the University of California, San Francisco Committee of Human Research (11-05995).

### Study Cohorts

The controlled Human Malaria Infection (CHMI) studies were conducted at the University of Sunshine Coast Clinical Trials Unit or Q-Pharm in Brisbane, Australia, from 2012 to 2023. Inoculum preparation, volunteer recruitment, infection, monitoring and treatment were performed as previously described ^35,42,43,49^. Healthy adults (not pregnant or lactating) between 18 and 45-55 years and with no prior malaria exposure were eligible to participate. Participants were intravenously infected with 2800 viable *P. falciparum* infected RBC (3D7 strain) and parasitemia monitored using quantitative polymerase chain reaction (qPCR)^62^. Participants were treated with the anti-malarial drugs 8-9 days post-infection. Blood samples were collected in lithium heparin tubes at days 0 (before infection, I), 8/9 (prior to anti-malarial treatment, T), 15/16 (peak T cell activation, T+7) and 29-42 (convalescence, T+29-42). Samples used here were collected from volunteers who consented to donate blood for immunological studies within the parent clinical trials of the following studies NCT02867059^35^, ACTRN1262-0000995976^42^, NCT03542149^43^, NCT05979207, ACTRN12621-000866808^49^ (Table S1). Additionally, to assess the impact of type I IFN signaling blockade and responses in second infection, samples from participants from a randomised, double-blind, placebo-controlled trial to evaluate ruxolitinib co-administered with artemether-lumefantrine were used (ACTRN12621-000866808) ^49^ (Table S1). For all studies, PBMCs were isolated by Ficoll Paque Plus (Sigma Aldrich; Cat #GE17- 1440-03) density centrifugation. Additionally, for *in vitro* malaria-naive healthy control assays, PBMCs were collected from healthy non-infected malaria naïve donors and from volunteers’ blood donation to the Australian Red Cross Lifeblood^TM^ service in Brisbane or Melbourne in Australia, with the same isolation method. Isolated PBMCs were cryopreserved in 10% DMSO/FCS at concentrations of 10^7^ cells/mL. For all assays, cryopreserved PBMCs were thawed at 37°C and after two washes in Roswell Park Memorial Institute 1640 media (RPMI, containing L- Glutamine, 25nM HEPES; Gibco-BRL; Cat #11875093) containing 10% FCS (R10) then RPMI medium alone, were counted using a DeNovix CellDrop FL Cell Counter, spun down and aliquoted to concentrations of 10^7^ cells/mL in 10% FCS/RPMI. Pooled plasma from Ugandan individuals with high malaria exposure was obtained from participants enrolled in a longitudinal study by the East African International Centers of Excellence in Malaria Research conducted in the high transmission areas of Uganda^63,64^.

### 3D7-Plasmodium *falciparum* culture

Packed red blood cells (RBCs) from O negative blood donors were infected in vitro with the *Pf* - 3D7 parasite strain^65^. Packed RBCs for parasite culture were acquired from the Australian Red Cross. *Pf*-infected RBCs (pRBCs) were cultured at 5% haematocrit in RPMI supplemented with AlbuMAX II (0.25%) and heat-inactivated human sera (5%). Cultures were incubated at 37°C in 1% O_2_, 5% CO_2_, 94% N gas mixture. Culture media was replaced daily, and Giemsa-stained blood smears monitored parasite stage/parasitemia. Synchronised parasite cultures were grown to 15% parasitemia then purified via magnet separation to enrich mature trophozoite stage pRBCs. Approximately 10^8^ purified pRBCs per vial (>95% purity) were stored at -80°C after adding a Glycerolyte cryopreservant at a 2:1 ratio. Alongside, 300ul of packed autologous uninfected red blood cells (uRBCs) were likewise cryopreserved for controls used in each assay. Aliquots of magnet purified pRBCs and uRBCs were also freeze/thawed 5x, sonicated, then centrifuged and supernatant collected the isolate cell lysate.

### Single-cell RNA sequencing of γδ T cells during first infection

#### Study participants and peripheral blood mononuclear cell processing

To investigate malaria-driven transcriptional changes in γδ T cells during primary infection by scRNAseq, we sequenced isolated γδ T cells from CHMI participants (NCT02867059^35^, Table S1) pre-infection (day 0) and at 8-, 16- and 36-days post-infection. PBMCs were thawed and then stained *ex vivo* to sort live γδ T cells (Table S4). PBMCs were washed with 2% FCS/PBS and then surface stained with anti-TCR γδ FITC for 15 minutes, then remaining surface antibodies were added for an additional 15 minutes. PBMCs were stained for viability with Sytox Blue and live γδ T cells were sorted on BD FACSAria^TM^ III Cell Sorter into 2% FCS/PBS (Fig. S1A). Sorted γδ T cells purity was approximately 92%.

#### *10X* Genomics Chromium GEX Library preparation and sequencing

Sorted cells γδ T cells from CHMI participants (*n* = 4, 4 timepoints) were pooled and loaded into each lane of Chromium Next GEM Single Cell 5’ Reagent Kit v2 and Gel Bead-in- Emulsion (GEMs) generated in the 10X Chromium Controller (Chromium Next GEM Single Cell 5’ Kit v2, PN-1000263, Chromium Next GEM Chip K Single Cell Kit, PN-1000286, Library Construction Kit, PN-1000190, Dual Index Kit TT Set A, PN-1000215). 5’ Gene Expression Libraries were generated according to the manufacturer’s instructions. Generated libraries were sequenced in a NextSeq 550 System using High Output Kit (150 Cycles) version 1 according to the manufacturer’s protocol using paired-end sequencing (150-bp Read 1 and 150 bp Read 2) with the following parameters Read 1: 26 cycles, i7 index: 10 cycles, i5 index: 10 cycles and Read 2: 90 cycles.

#### Single-cell RNAseq transcriptomic analysis

ScRNAseq data was demultiplexed, aligned and quantified using Cell Ranger v3.1.0 (10x Genomics) against the human reference genome (GRCh38-3.0.0), with default parameters. Cell genotype was obtained using cellsnp-lite^66^ with the list of informative SNPs obtained from 1000_Genome_Project^67^. Then, we use vireo^68^ to predict the original donor utilising the cell’s haplotype ratios. Cell ranger count matrices for each sample (donor and day) were loaded, merged and analyzed using Seurat package v4^69^. Cell cycle scores, mitochondria DNA transcripts and total/unique gene counts were calculated per cell using Seurat built-in functions. Cell cycle score was assigned to each cell using the CellCycleScoring function and evaluated with Principal Component Analysis (PCA). Cells with less than 7.5% of mitochondria DNA transcripts and between 700 and 2700 genes per cell were retained (Fig. S1B). Contaminating monocytes from the sorting process were detected by RNA transcripts and removed.

Haemoglobin-associated and TRAV/TRBV gene expression were removed before clustering. The filtered dataset was scaled to regress the effects of mitochondria DNA transcripts content and cell cycle using the ScaleData function, which regresses each variable individually. Filtered data was split per donor and day, then each dataset was normalised with SCT transform v2^70^, and the most variable genes in each were selected using FindVariableFeatures. Before integration, the most variable genes shared among the datasets were identified using FindIntegrationAnchors and used to integrate the datasets using the IntegrateData function. During integration, we also performed CellCycleScoring to improve the accuracy and robustness of downstream analyzes by accounting for the effects of the cell cycle on gene expression. Integration by this method removed variation from differences between donors and days (Fig. S1C).

#### Cell clustering

We applied a linear transformation using ScaleData and regressed out mitochondrial contamination, a source of unwanted variation. The top 30 principal components (PCs) were calculated using RunPCA. From these 30 PCs, we constructed a k-nearest neigh- bour graph Euclidean distance in PCA space and refined the edge weights between any two cells based on the shared overlap in their local neighbourhoods (Jaccard similarity) based on standard Seurat workflow. Next, we applied the Louvain algorithm as our modularity optimisation technique to set the granularity of clusters at a resolution of 0.6. The non-linear dimensionality reduction technique ‘UMAP’ visualised clusters in 2D space with 13 dimensions. Clusters were annotated based on enriched genes identified with the FindAllMarkers function. Cluster frequencies are compared A Wald-test-based linear mixed-effects model analysis of cluster proportions across timepoints with donor-level random effects, followed by global FDR correction.

#### Differential gene expression analysis

Longitudinal differential gene expression from day 0 (baseline) was compared to baseline levels using a pseudobulk approach that controls for false discoveries introduced by inter-sample variability and low cell numbers^71,72^. The normalized count matrix and the metadata were extracted from the Seurat object. Counts were aggregated per sample (i.e., donor and day) for each cluster. Significant differentially expressed genes between each post-infection days and baseline (day 0) were determined using the quasi-likelihood *F* test with an FDR < 0.05 using edgeR^73^. Shared and subset-specific DEGs were visualised using Upset plots using the UpSetR package v1.4.0^74^. Clusters were annotated based on previous shown in previous literature^14,38,39,75^.

Pathway analysis was done using the SCPA R package^41^. Raw counts were extracted from the SCT assay, and cells were grouped by annotated cell types and timepoints. Cells were stratified by cell type and day, and subsets were extracted using the seurat_extract function within SCPA package. Pathway gene sets were obtained from the Molecular Signatures Database using the msigdbr package^76^ and filtered to retain pathways containing 15–200 genes. Pathway comparisons between timepoints (day 0 vs day 8, day 0 vs day 36, and multi-group comparisons) were performed using compare_pathways with default parameters. This approach evaluates pathway-level significance based on coordinated gene expression changes rather than predefined gene lists. Results were summarised as pathway-level −log q-values and fold changes.

### Immunophenotyping of flow cytometry data

In all instances, flow cytometry data was generated from PBMCs cryopreserved and thawed as above, except for one instance (Phenotyping 1, Table S5). 10^6^ PBMCs were washed with 2% FCS/PBS, and viability staining was performed for 15 minutes with viability dye. Washed cells were surface stained with TCR γδ PerCPCy5.5 for 15 minutes then remaining surface antibodies were added for an additional 15 minutes. Washed cells were fixed and permeabilized with eBioscience^TM^ Foxp3 / Transcription Factor Staining Buffer Set (Thermo Fisher Scientific (MA, USA); Cat. #00-5523-00) for 20 minutes on ice, washed, then intracellular staining was performed for 30 minutes with fluorescent-tagged antibodies. Washed cells were re-suspended in 2% FCS/PBS and events collected with flow cytometer. Specific cohorts and staining information detailed in Supplementary Table S1, S5-11.

### *In vitro* parasite stimulation and cytokine capture

PBMCs from CHMI participants (*n* = 6, ACTRN1262-0000995976^42^, Table S1) were thawed, then rested for 2 hrs in R10 at 37°C. Live mid-late trophozoite-stage *Pf* was isolated by magnet purification at a purity ≥ 99%. 10^7^/mL RBCs were stained with Cell Trace Violet (1/2500; Biolegend #CD34557A) for 20 minutes in a 37°C water bath, quenched with R10 for 10 minutes then washed with R10 twice. Trophozoite-stage *Pf* were opsonized with hyperimmune plasma from Ugandan children in the PRISM cohort^63^, or CHMI participant- and day-matched plasma corresponding to PBMC collections for 1 hour at 37°C in serum-free media. 10^6^ PBMCs were combined in culture for 8 hours at 37°C with unopsonized pRBC, pRBC opsonized with PRISM plasma, pRBC opsonized with CHMI plasma and uRBC. Brefeldin A (10mg/mL; BD Biosciences Cat. #555029), monensin (10mg/mL; BD Biosciences; Cat. #554274) and CD107a BUV395 (1/200; Invitrogen #565113) were added at the beginning of co-cultures. After co-culture, cells were immunophenotyped as above.

### *In vitro* APC induction

For induction of T cell activating surface protein expression by γδ T cells after in vitro culture, 10^6^ malaria-naïve PBMCs (*n* = 15, ARC) were co-cultured with blank media, 100nM HMBPP, 10^6^ uRBCs or 10^6^ pRBCs for 5 days at 37°C, 5% CO_2_ in R10 supplemented with 100 IU/mL IL-2 (Miltenyi Biotec; Cat. #130-097-745). Co-cultured PBMCs were immunophenotyped as above.

### Mixed lymphocyte reaction

For the mixed lymphocyte reaction assay of γδ and naive CD4 T cells, cells were isolated from PBMCs using the TCRγ/δ+ T Cell Isolation Kit (Miltenyi Biotec; Cat. #130-092-892), Naive CD4+ T Cell Isolation Kit II (Miltenyi Biotec; Cat. #130- 094-131), a QuadroMACST M Separator (Miltenyi Biotec; Cat. #130-090-976), pre-Separation Filters (Miltenyi Biotec; Cat. #130-041-407) and LS Colums (Miltenyi Biotec; Cat. #130-042- 401) as per manufacturer’s instructions. γδ T cells were isolated from malaria-naïve PBMCs (ARC) co-cultured with blank media, 10^6^ uRBCs or 10^6^ pRBCs for 3 days (ensure high effector cell viability) at 37°C, 5% CO_2_ in R10. Matching γδ T and allogenic naive CD4 T cells were isolated from freshly thawed PBMCs. Isolated cells were counted using a DeNovix CellDrop^TM^ FL, spun down and re-suspended to concentrations of 10^7^ cells/mL in R10. Resultant effector and target cell populations were phenotyped prior to mixed lymphocyte reaction (Fig S8A-D, Table S9). Sorted naive CD4^+^ T cells were labelled with 5 μM of CellTrace Violet (1/2500; Biolegend #CD34557A) in PBS for 20 minutes in a 37°C, quenched for 10 minutes in R10 before being used as responder cells in the mixed lymphocyte reaction. 5 x 10^4^ isolated naive CD4 T cells (*n* = 12) were combined in an allogenic mixed lymphocyte reaction with 10^5^ isolated γδ T cells (*n* = 6) *ex vivo*, and from media, uRBC or pRBC cultures, and with Human T-Activator CD3/CD28 DynabeadsTM (Gibco, Cat. #11161D) plus recombinant cytokines 1ng/mL IL-12 (Miltenyi Biotec; Cat. #130-096- 704), 25ng/mL IL-23 (Miltenyi Biotec; Cat. #130-095-757) and 5ng/mL TGF-β1(Miltenyi Biotec; #130-095-067) for 6 days at 37°C, 5% CO2 R10 supplemented with 100 IU/mL IL-2^77^. After 6-days co-culture, cells were immunophenotyped as above

### *In vitro* cytokine activation

PBMCS from malaria-naïve donors (*n* = 6, HDC) were thawed as above. 10^6^ PBMCs were combined in culture for 19 hours at 37°C with 1:1 pRBC lysate or blank media, alone or with CXCL9 (50ng/mL; R&D Systems Cat #392-MG-010) and CXCL10 (50ng/mL; R&D Systems Cat #226-IP-010) or IL-12 (50ng/mL; R&D Systems Cat #219-IL-025), IL-15 (10ng/mL; ThermoFisher Scientific Cat #200-15-2UG), and IL-18 (250ng/mL; R&D Systems Cat #9124-IL-050/CF) in R10. Brefeldin A (10mg/mL; BD Biosciences Cat. #555029), monensin (10mg/mL; BD Biosciences; Cat. #554274) and CD107a BUV395 (1/200; Invitrogen #565113) were added at the beginning of co-cultures. To confirm, PBMCS from additional malaria-naïve donors HDC, n=6) were thawed and stimulated as above. In addition, we incubated PBMCs with mab 103.2 (100ug/mL, BTN3A blocking antibody^78^), a mouse anti-human IgG3 Antibody control (100ug/mL; Invitrogen #05-3600) or no mab for 30 minutes at 37°C prior to stimulation. We also included single stimulation conditions of with IL-12, IL-15 or IL-18. After co-culture, cells were immunophenotyped as above.

### ATAC-seq

#### Cell isolation

PBMCs from participants enrolled in CHMI studies (*n* = 6, ACTRN1262-0000995976^42^ and NCT05979207, Table S1) from 0, 8, 15, and 27/29/35 days post *Pf*-infection were thawed as above and pelleted cells resuspended in 25μL Brilliant Stain Buffer (BD) containing 1:20 Fc Block (Human Trustain FCX, Biolegend, 422301) and 1:500 viability dye (eflour 506, Thermo Fisher, 65-0866-14) for 10 minutes prior to the addition of 25μL surface staining cocktail in FACS buffer (0.1% BSA, 2mM EDTA in PBS) (Table S12). Following 30 minutes incubation on ice, cells were washed with 5mL FACS buffer, centrifuged (300 x g for 5 minutes at 4°C) and resuspended in 500 mL FACS buffer for sorting. Gating was performed to isolate Vδ2 T cells with the BD FACSAria Fusion (Fig. S14A).

#### ATAC-seq transposition

Vδ2 T cells were transposed for ATAC-seq using the omni-ATAC method^79,80^. Briefly, 50,000 cells were centrifuged (500 x g) and thoroughly resuspended in 50μL cold lysis buffer (ATAC Resuspension Buffer containing 0.1% Tween-20, 0.1% NP40, and 0.01% Digitonin) before 3 minutes incubation on an ice block. Samples were washed out with 450μL ATAC Resuspension buffer with 0.1% Tween-20, and nuclei pelleted via centrifugation (600 x g for 10 min at 4°C) prior to addition of 40μL freshly prepared transposition mixture (containing 2μL Tn5 transposase, 0.01% Digitonin and 0.1% Tween-20). Samples were incubated at 37°C for 40 minutes in a Thermomixer (Eppendorf; 1000 RPM). Following transposition, 250μL DNA binding buffer from the Clean and Concentrator-5 kit (Zymo, D4014) was added and reactions stored at -80°C.

#### Library preparation and sequencing

Transposed DNA was cleaned using the Clean and Concentrator-5 kit (Zymo, D4014) according to the manufacturer’s instructions and eluted in 21μL of elution buffer. DNA was subjected to PCR amplification (72°C 5 min, 98°C 30 sec, then 8-15 cycles of 98°C 10 sec, 63°C 30 sec and 72°C 1 min) with adaptor-specific primers containing Illumina index barcode sequences, as previously described^79,80^. Amplified libraries were cleaned up using Zymo DNA Clean and Concentrator 5 columns followed by RHS clean-up with SPRIselect beads (Beckman Coulter) to deplete large molecular weight fragments >1000bp. For pre-sequencing quality control (QC), PCR was performed on libraries to assess enrichment of accessible regions, selected manually from publicly available ATAC-seq data.

qPCR was performed using 5μL of 1:10 diluted library using the PowerUp SYBR Green Master Mix (Applied Biosystems) according to the manufacturer’s instructions. Genomic DNA (gDNA; 1ng/μL) was run as a comparison. The accessibility ratio was calculated as the mean copy number of accessible regions divided by the mean copy number of inaccessible regions, relative to gDNA. Enrichment was >1 for all samples. Libraries were visualised using a Tapestation instrument (Agilent) using the High Sensitivity DNA Assay (Agilent) kit, according to the manufacturer’s instructions, to confirm that fragment sizes were consistent with high-quality ATAC-seq libraries, and to determine library concentration. Following QC, libraries were sequenced on a MGI DNBSEQ T7 Flow Cell (2x150bp reads).

#### ATAC-seq transcriptomic analysis

Fastq read quality was assessed using FastQC version v0.12.1 and summarized with MultiQC^81^. Fastq files were processed using the ENCODE pipeline (v1.5.4)^82^. Briefly, adapter sequences were removed using Cutadapt v1.9.1. Reads were then aligned to the GRCh38 human genome using Bowtie v2.2.6 “–very sensitive” mode^83^. SAMtools v1.7 was used to convert to BAM format and sort alignments^84^. The Bioconductor package ATACseqQC (v1.30.0) was used to assess sample quality and library complexity^85^. Low quality reads were removed (MAPQ<30) and the fraction of mithocondrial reads was calculated using (SAMstats v.0.21). PCR duplicates were labelled and removed using Picard v1.126. Peaks were called across all samples using MACS2v2(2.1.0) with a p value threshold of 0.01, excluding mitochondrial peaks. Bedtools v 2.31.1 was used to include the peaks present in at least 4 of the 21 samples. Feature Counts v2.0.6) was used to count aligned reads to generate a count per sample for each peak ^86^. These counts were then imported into R v4.4.0 for further statistical analysis. Peaks were annotated with ChIPseeker v1.42.1. Surrogate variable analysis was conducted to capture batch effect and difference in samples quality in our model using the sva package (v.3.54.0)^87^. Limma (v.3.62.2)^88^ and variancePartition (v.1.36.3)^89^ were used to normalize the data (using trimmed mean of M-values method) and perform differential accessibility analyzes (with the dream function) including individuals as random effect (∼ day + sv + (1|samples)^90^. QC metrics (Table S12) were assessed as described in ENCODE consortium guidelines (Available at https://www.encodeproject.org/atac-seq/), including sequencing depth (100% of samples had >22 million paired-end reads), alignment rate (all samples had alignment rates of >98% except for 2 with 61% and 72%) transcription start site enrichment (2 samples had ‘concerning’ scores “<5”, 1 was ‘acceptable’ “5-7” and the remaining 18 were ‘ideal’>7) and, fraction of reads in called peaks (FRiP score; 100% of samples >0.3). Libraries complexity post PCR duplicate removal showed an "ideal” non-redundant fraction (NRF) and PCR bottlenecking coefficients. Results pre duplicate removal are shown for transparency in (Table S12). ATAC-seq data were of acceptable quality based on sequencing depth and enrichment of reads near transitional start sites (Table S13).

### Statistical Analysis

Cell events collected by flow cytometry were analyzed using BD Flowjo v10.7. Statistical analysis of cell frequencies was performed using R (v4.0.3) packages’ ggplot2’ (v3.4.1) and ‘ggpubr’ (v0.4.0). All statistical tests were two-sided. Frequencies and proportions of cell responses were compared with the Wilcoxon signed rank test, for paired observations, and the Mann-Whitney U test for unpaired observations. P values ≤ 0.05 were considered significant.

Linear mixed effects models (LMMs) were used to calculate changes in γδ T cell frequencies between longitudinal timepoint measurements and between ruxolitinib and placebo treatment groups^49^. The lmer function of the lme4 R package v1.1-34 was used to fit the model with the restricted maximum likelihood (REML) option selected. Cell frequencies of γδ T cell subsets generated from flow cytometry were fit as outcome variables, while sampling timepoints and treatment groups were fit as interacting fixed effects. The individual identifier was fit as a random effect to account for repeated measurements. Outcome variables were pre-processed before analysis: 1) log10 normalised and 2) zero values were transformed to minimum non-zero values divided by two. Residual bootstrapping and multiple comparisons adjustment were applied as previously indicated ^49^.

A segmented regression approach was used to assess changes in inflammatory and immune responses from the infection to treatment time point and how these changes differed between first and second infection (phase) and between treatment. Log-linear mixed-effect models included day, timepoint, treatment, phase, and a phase-by-day interaction as fixed effects, with participant-level random effect and REML estimation. Residual bootstrapping and multiple comparisons adjustment were applied as previously indicated published^49^. Immunology analysis was conducted using R version 4.3.3.

## Supporting information

Supplementary_Tables_ext

## Data availability

Fastq files, and raw data files for the scRNAseq analysis can be found under NCBI accession number GSE253661 and seurat object here: **10.5281/zenodo.19673743.**

## Acknowledgements

The Burnet Institute and QIMR Berghofer acknowledge the Traditional Owners and custodians of the lands where they are located, Boonwurrung people of the Kulin nations and the Turrbal and Yugara people. We acknowledge and thank the study participants and the staff at University of Sunshine Coast and Q-Pharm Clinical Trials Units, and staff at the Queensland Paediatric Infectious Diseases Laboratory for conducting the malaria PCR assays. We thank staff at the Clinical Malaria Group, QIMR-Berghofer, for inoculum preparation and assistance with managing the clinical trials. We also thank the South Australian Genomics Center (SAGC) for help with ATAC sequencing. The SAGC is supported by the National Collaborative Research Infrastructure Strategy (NCRIS) through Bioplatforms Australia and by the SAGC partner institutes. We acknowledge Dr. Katherine O’Flaherty for provision of pRBC lysate used to optimize experimental protocol for data shown in Figure 6B-D and S13. We thank Dr Adam Ulrich for providing BTN3A blocking antibody. We thank Lifeblood Biological Resources Australia for providing the human red blood cells and buffy coats.

## Author contributions

N.L.D. and M.J.B. conceptualized the study;

N.L.D, D.A., T.F., J.R.L., F.R., R.M., J.L., L.B., J.E., and N.E.S., generated data, supervised by

M.J.B, C.E, J.AL, F.J.R. and D.J.L

N.L.D, Z.P, D.O analysed data

R.W, J.S.M., B.E.B, M.K, and I.S, conduced and managed clinical studies, and provided study samples

J.M., B.E.B., CE, and M.J.B provided funding

N.L.D. and M.J.B. wrote the manuscript, with feedback and input from all authors.

