## Supplementary_Tables_Figures for "Vδ2 T cell activation by malaria is enhanced in second infection via the cell extrinsic cytokine milieu"

**Supplementary Tables and Figures**

***Supplementary Table S1****: Cohort characteristics*

| **Cohort** | **Study ID** | **n** | **Age, median [IQR] (y)** | **Male (%)** | **Assay** | **Cytometer** | **References** |
| --- | --- | --- | --- | --- | --- | --- | --- |
| scRNA-seq sort | NCT02867059 | 4 | 28.5 [25.5–35] | 50 | Surface + sort | BD FACSAriaIII | Fig. 1/2, Fig. S1/2;  Table S2-4; |
| Phenotyping 1 | ACTRN1262-0000995976 | 12 | 29.9 [22.1–37.7] | 50 | Whole blood fixed (PROT1) surface | Cytek Aurora (5L) | Fig. 3A-C, 4A S3A/B, S6; Table S5 |
| Phenotyping 2 | NCT03542149 | 8 | 22.5 [20.0–23.5] | 62.5 | Surface + intracellular | Cytek Aurora (5L) | Fig. 3D/E, S3C/D  Table S6; Fig. |
| Parasite stimulation + cytokine capture | CHMI (ACTRN1262-0000995976) | 6 | 25.6 [22.7–31.5] | 67 | Surface + intracellular | Cytek Aurora (5L) | Fig. 3F/G, 4D, S4, S5, S9;  Table S7 |
| APC induction | ARC malaria-naïve donors | 15 | – | – | Surface | BD LSR Fortessa (5L) | Fig 4B, S7; Table S8 |
| MLR (purity control) | ARC malaria-naïve donors | 7 | – | – | Surface | BD LSR Fortessa (5L) | Fig. S8A-D; Table S9 |
| MLR | ARC malaria-naïve donors | 12 | – | – | Surface | BD LSR Fortessa (5L) | Fig. 4C, S8E; Table S10 |
| 1^st^ Infection: Ruxolitinib | ACTRN12621-000866808 | 11 | 38 [31–44] | 64 | Surface + intracellular | Cytek Aurora (5L) | Fig. 5, 6A, 7, S10-12, S15;  Table S6 |
| 1^st^ Infection: Placebo | ACTRN12621-000866808 | 9 | 27 [23–34] | 44 |  |  |  |
| 2^nd^ Infection: Ruxolitinib | ACTRN12621-000866808 | 9 | 38 [32–41] | 56 |  |  |  |
| 2^nd^ Infection: Placebo | ACTRN12621-000866808 | 6 | 28 [27–34] | 33 |  |  |  |
| Cytokine activation | HDC malaria-naïve donors | 12 | – | – | Surface + intracellular | Cytek Aurora (5L) | Fig. 6B-D S13; Table S11 |
| ATAC-seq sort | ACTRN1262-0000995976; NCT05979207 | 6 | 25.0 [20.0–36.0] | 50 | Surface + sort | BD FACSAriaIII | Fig. S14; Table S12-14 |

***Supplementary Table S2:*** *DEG of cluster annotation –* attached as excel

***Supplementary Table S3****: DEG for longitudinal analysis during CHMI –* attached as excel

***Supplementary Table S4****: scRNAseq sort panel*

| **Antigen** | **Fluorophore** | **Clone** | **Manufacturer** | **Cat #** | **Concentration** |
| --- | --- | --- | --- | --- | --- |
| Surface | | | | | |
| TCR γδ | FITC | B1 | Biolegend | 331208 | 3/25 |
| CD19 | PE | HIB19 | Biolegend | 302208 | 1/100 |
| TCR Vδ2 | APC | B6 | Biolegend | 331418 | 1/50 |
| CD3 | AF700 | SK7 | Biolegend | 344822 | 1/50 |
| Viability | Sytox Blue |  | Invitrogen | S34857 | 1/20 |

***Supplementary Table S5****: Phenotyping 1 panel*

| **Antigen** | **Fluorophore** | **Clone** | **Manufacturer** | **Cat #** | **Concentration** |
| --- | --- | --- | --- | --- | --- |
| Surface | | | | | |
| TCR Vδ1 | APC | TS8.2 | Invitrogen | 17-5679-42 | 1/100 |
| TCR Vδ2 | APC-FIRE | B6 | Biolegend | 331418 | 1/100 |
| CD19 | BUV395 | SJ25C1 | BD | 563549 | 1/100 |
| CD16 | BUV737 | 3G8 | BD | 612787 | 1/200 |
| CD3 | BUV805 | SK7 | BD | 612893 | 1/200 |
| CD27 | BV421 | M-T271 | BD | 562513 | 2/50 |
| CD38 | BV480 | HIT2 | BD | 566137 | 1/500 |
| CD86 | PE-Cy7 | 2331 (FUN-1) | BD | 561128 | 1/100 |
| CD45RA | BUV563 | HI100 | BD | 565702 | 1/1000 |
| ICOS | BV650 | DX29 | BD | 563832 | 1/50 |
| HLADR | BV750 | L243 | Biolegend | 307672 | 1/50 |
| CD98 | FITC | MEM-108 | Biolegend | 315603 | 1/50 |

***Supplementary Table S6****: Phenotyping 2 panel*

| **Marker** | **Fluorophore** | **Clone** | **Manufacturer** | **Cat #** | **Concentration** |
| --- | --- | --- | --- | --- | --- |
| Viability | | | | | |
| Viability | Live Dead Blue | - | Invitrogen | L34962A | 1/2500 |
| Surface 1 | | | | | |
| TCR γδ | PerCPCy5.5 | B1 | BD | 564157 | 3/25 |
| Surface 2 | | | | | |
| CD16 | BUV395 | 3G8 | BD | 563785 | 1/400 |
| PD1 | BUV615 | EH12.1 | BD | 612991 | 1/50 |
| CD56 | BUV737 | NCAM16.2 | BD | 612766 | 1/400 |
| CD3 | BUV805 | SK7 | BD | 612893 | 1/400 |
| CD27 | BV421 | M-T271 | Biolegend | 356418 | 1/200 |
| CD57 | Pacific Blue | HNK-1 | Biolegend | 359607 | 1/200 |
| CD14 | BV510 | M5E2 | Biolegend | 301842 | 1/50 |
| CD19 | BV510 | SJ25C1 | Biolegend | 363020 | 1/50 |
| CD127 | BV570 | A019D5 | Biolegend | 351308 | 3/50 |
| CD38 | BV605 | HB7 | BD | 562665 | 1/200 |
| CD161 | BV650 | DX12 | BD | 563864 | 1/20 |
| CD45RA | BV711 | HI100 | Biolegend | 304128 | 1/400 |
| HLA-DR | BV750 | L243 | Biolegend | 307672 | 1/100 |
| CD8 | BV785 | SK1 | Biolegend | 344740 | 1/400 |
| KLRG1 | PE-Cy7 | 13F12F2 | Invitrogen | 25-9488-42 | 1/200 |
| TCR Vδ2 | APC-Fire 750 | B6 | Biolegend | 331420 | 1/100 |
| Intracellular | | | | | |
| Granulysin | AF488 | RB1 | BD | 558254 | 3/50 |
| NKG7 | PE | 2G9A10F5 | Beckman-Coulter | IM3293 | 3/5000 |
| Perforin | PE-CF594 | (delta)G9 | BD | 563763 | 1/10000 |
| Granzyme B | APC | QA16A02 | Biolegend | 372204 | 3/5000 |

***Supplementary Table S7****: In vitro cytotoxicity/phagocytosis panel*

| **Antigen** | **Fluorophore** | **Clone** | **Manufacturer** | **Cat #** | **Concentration** |
| --- | --- | --- | --- | --- | --- |
| Viability | | | | | |
| Viability | Live Dead Blue | - | Invitrogen | L34962A | 1/2500 |
| Surface 1 | | | | | |
| TCR γδ | PerCPCy5.5 | B1 | Invitrogen | 564157 | 5/100 |
| Surface 2 | | | | | |
| CD3 | BUV805 | SK7 | BD | 612893 | 1/400 |
| CD19 | BV510 | SJ25C1 | BD | 363020 | 1/50 |
| CD14 | BV650 | MSE2 | Biolegend | 301836 | 1/200 |
| CD16 | AF700 | 3G8 | Biolegend | 302026 | 1/500 |
| TCR Vδ2 | APC/Fire 750 | B6 | Biolegend | 331420 | 1/100 |
| Intracellular | | | | | |
| NKG7 | PE | 2G9A10F5 | Biolegend | IM3293 | 1/2500 |
| Granulysin | AF488 | RB1 | Beckman-Coulter | 558254 | 2/25 |
| Perforin | PE-CF594 | (delta)G9 | BD | 563763 | 3/2500 |
| Granzyme B | APC | QA16A02 | BD | 372203 | 3/5000 |
| TNF | BV750 | Mab11 | Biolegend | 566359 | 1/100 |
| IFNγ | BV605 | B27 | BD | 562974 | 1/50 |

***Supplementary Table S8****:* In vitro *APC induc*tion *panel*

| **Antigen** | **Fluorophore** | **Clone** | **Manufacturer** | **Cat #** | **Concentration** |
| --- | --- | --- | --- | --- | --- |
| Viability | | | | | |
| Viability | Zombie Aqua | - | Biolegend | 423101 | 1/500 |
| Surface | | | | | |
| TCR Vδ1 | APC | TS8.2 | Invitrogen | 17-5679-42 | 1/100 |
| TCR Vδ2 | APC-FIRE | B6 | Biolegend | 331418 | 1/100 |
| CD3 | BUV737 | UCHT1 | BD | 612750 | 3/100 |
| CD14 | BV510 | M5E2 | Biolegend | 301842 | 1/50 |
| CD19 | BV510 | SJ25C1 | Biolegend | 363020 | 1/50 |
| CD40 | BV605 | 5C3 | Biolegend | 334336 | 1/50 |
| HLADR | BV785 | L243 | Biolegend | 307642 | 1/100 |
| CD86 | PE-Cy7 | 2331 (FUN-1) | BD | 561128 | 1/100 |

***Supplementary Table S9****: Mixed lymphocyte reaction purity panel*

| **Antigen** | **Fluorophore** | **Clone** | **Manufacturer** | **Cat #** | **Concentration** |
| --- | --- | --- | --- | --- | --- |
| Viability | | | | | |
| Viability | Live Dead NIR | - | Invitrogen | L34975 | 1/1000 |
| Surface | | | | | |
| CD3 | BUV737 | UCHT1 | BD | 612750 | 3/100 |
| CD45RA | BV650 | HI100 | BD | 304136 | 1/500 |
| HLADR | BV785 | L243 | Biolegend | 307642 | 1/100 |
| TCR Vδ1 | FITC | TS8.2 | Invitrogen | TCR2730 | 1/50 |
| TCR Vδ2 | APC | B6 | Biolegend | 331418 | 1/50 |
| CD4 | PerCP-Cy5.5 | OKT4 | Biolegend | 317428 | 1/50 |
| ICOS | PE | C398.4A | Biolegend | 313507 | 1/100 |

***Supplementary Table S10****: Mixed lymphocyte reaction culture antibody list*

| **Antigen** | **Fluorophore** | **Clone** | **Manufacturer** | **Cat #** | **Concentration** |
| --- | --- | --- | --- | --- | --- |
| Viability | | | | | |
| Viability | Live Dead NIR | - | Invitrogen | L34975 | 1/1000 |
| Surface | | | | | |
| TCR Vδ2 | APC | B6 | Biolegend | 331418 | 1/100 |
| CCR6 | BV650 | 11A9 | BD | 563922 | 1/50 |
| CXCR5 | BV711 | J252D4 | Biolegend | 356934 | 1/50 |
| CD4 | BV785 | OKT4 | Biolegend | 317442 | 3/100 |
| TCR Vδ1 | FITC | TS8.2 | Invitrogen | TCR2730 | 1/50 |
| ICOS | PE | C398.4A | Biolegend | 313507 | 3/500 |
| CXCR3 | PE-CF594 | 1C6/CXCR3 | BD | 562451 | 1/50 |
| PD1 | PE-CY7 | EH12.1 | BD | 561272 | 3/100 |
| CD3 | PerCP-Cy5.5 | OKT3 | Biolegend | 317335 | 3/100 |

***Supplementary Table S11****: In vitro cytokine activation panel*

| **Antigen** | **Fluorophore** | **Clone** | **Manufacturer** | **Cat #** | **Concentration** |
| --- | --- | --- | --- | --- | --- |
| Viability | | | | | |
| Viability | ViaDye Red | - | Cytek Biosciences | R7-60008 | 1/40000 |
| Surface 1 | | | | | |
| TCR γδ | PerCPCy5.5 | B1 | Invitrogen | 564157 | 5/100 |
| Surface 2 | | | | | |
| CD38 | BUV737 | HB7 | BD | 564686 | 1/200 |
| CD3 | BUV805 | SK7 | BD | 612893 | 1/200 |
| TCR Vδ2 | PE-Cy7 | B6 | Biolegend | 331422 | 1/100 |
| Intracellular | | | | | |
| TNF | BV750 | Mab11 | Biolegend | 566359 | 1/100 |
| IFNγ | BV605 | B27 | BD | 562974 | 1/50 |

***Supplementary Table S12****: ATAC-seq sort panel*

| **Antigen** | **Fluorophore** | **Clone** | **Manufacturer** | **Cat #** | **Concentration** |
| --- | --- | --- | --- | --- | --- |
| Viability | | | | | |
| Viability | eFluor506 | - | Invitrogen | 65-0866-14 | 1/500 |
| Surface | | | | | |
| CD19 | BV786 | SJ25-C1 | BD | 563325 | 1/50 |
| TCR Vδ2 | BV605 | B6 | Biolegend | 331531 | 1/50 |
| CD3 | FITC | HIT3α | BD | 561802 | 1/25 |

***Supplementary Table S13****: QC ATAC-seq –* attached as excel

***Supplementary Table S14****: DER ATAC-seq –* attached as excel

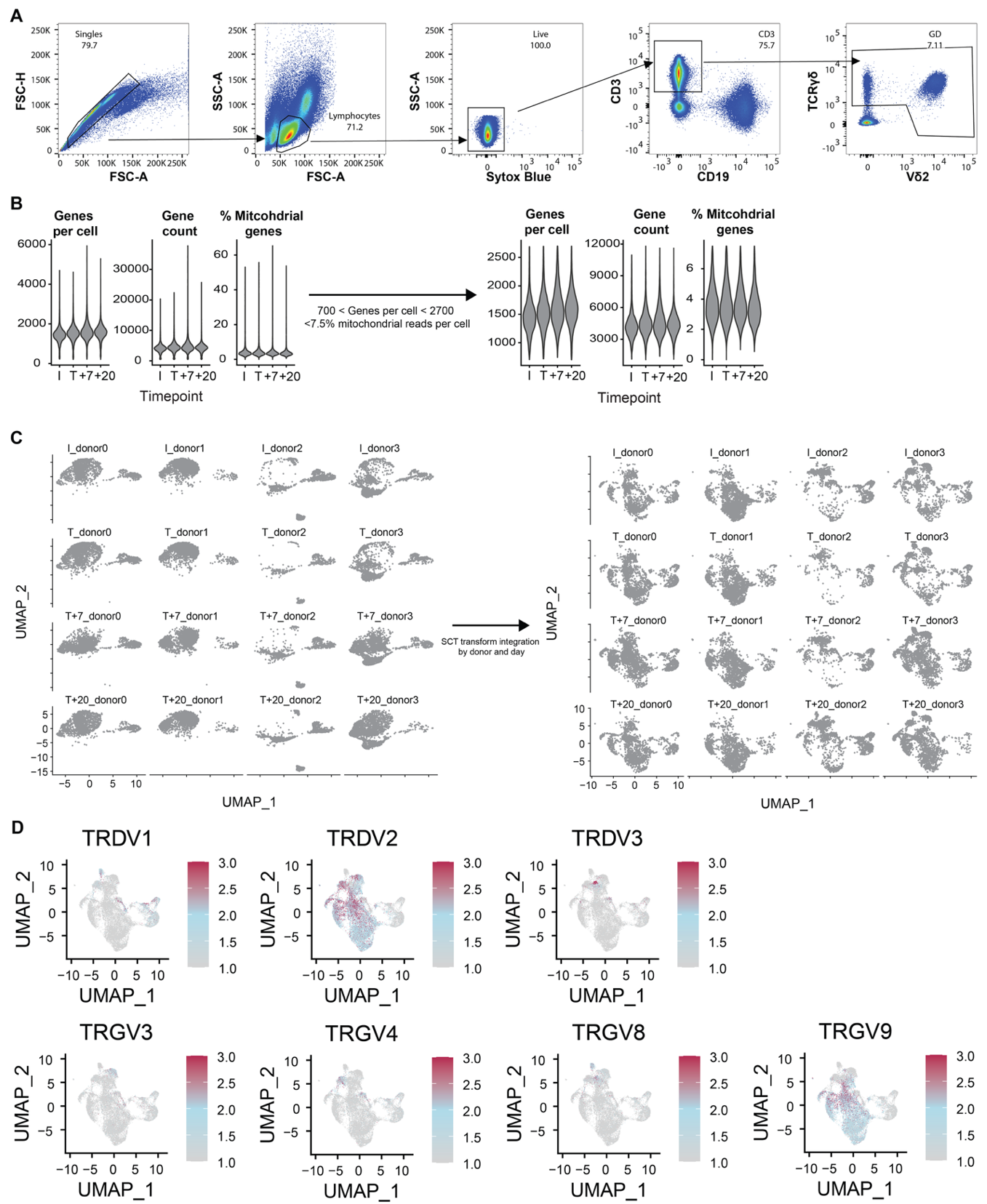

**Figure S1: Single cell RNAseq integration and TRDV/TRGV expression**

Participants (*n*=4) were intravenously infected with *Pf*-infected RBCs in a CHMI. Venous blood drawn at infection (I, 0 days post infection (d.p.i.)), treatment (I, 8 d.p.i.) and at 15 d.p.i. (T+7) and 36 d.p.i. (T+20). Single cell RNA sequencing libraries were constructed from isolated γδ T cells. (**A**) FACS gating strategy to purifiy γδ T cells. (**B**) Cells with greater than 7.5% of mitochondria DNA transcripts were removed, and between 700 and 2700 genes per cell were retained in the data set. (**C**) UMAP visualisation before and after scTransform data integration by donor and day. (**D**) UMAP visualisation of TRGV/TRDV expression. Related to Figure 1.

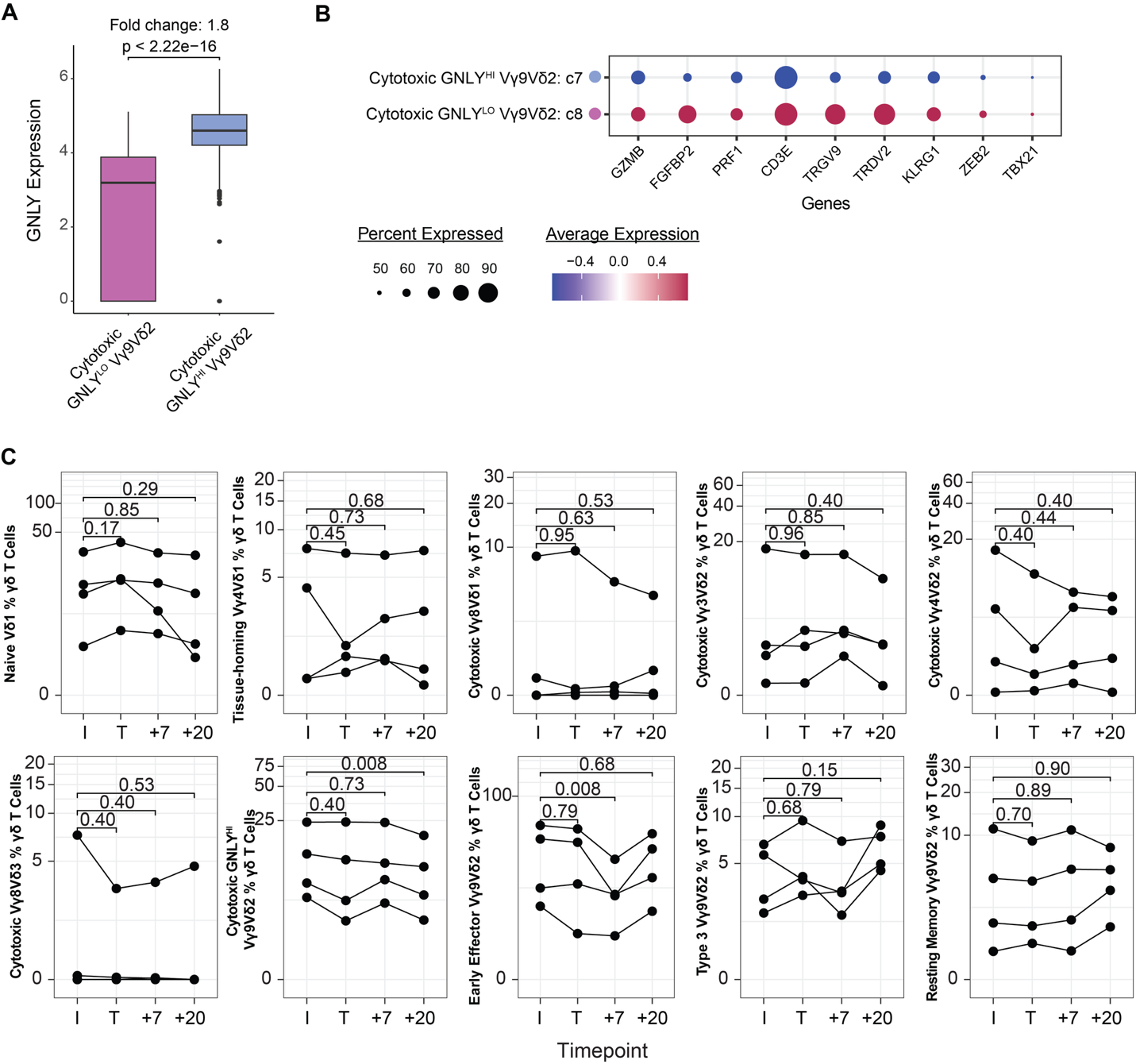

**Figure S2: scRNAseq clustering and longitudinal cluster proportions**

**A)** *GNLY* expression by *GNLY*^LO^ (c8) and *GNLY*^HI^ (c7) Vγ9Vδ2 T cells identified in scRNAseq dataset. P values from Mann-Whitney U test. (**B**) Dotplot of expression of marker genes in *GNLY*^LO^ (c8) and *GNLY*^HI^ (c7) Vγ9Vδ2 T cells. **C)** scRNAseq cluster proportions by donor across timepoints. P values from linear mixed effects model with FDR correction. Related to Figure 1.

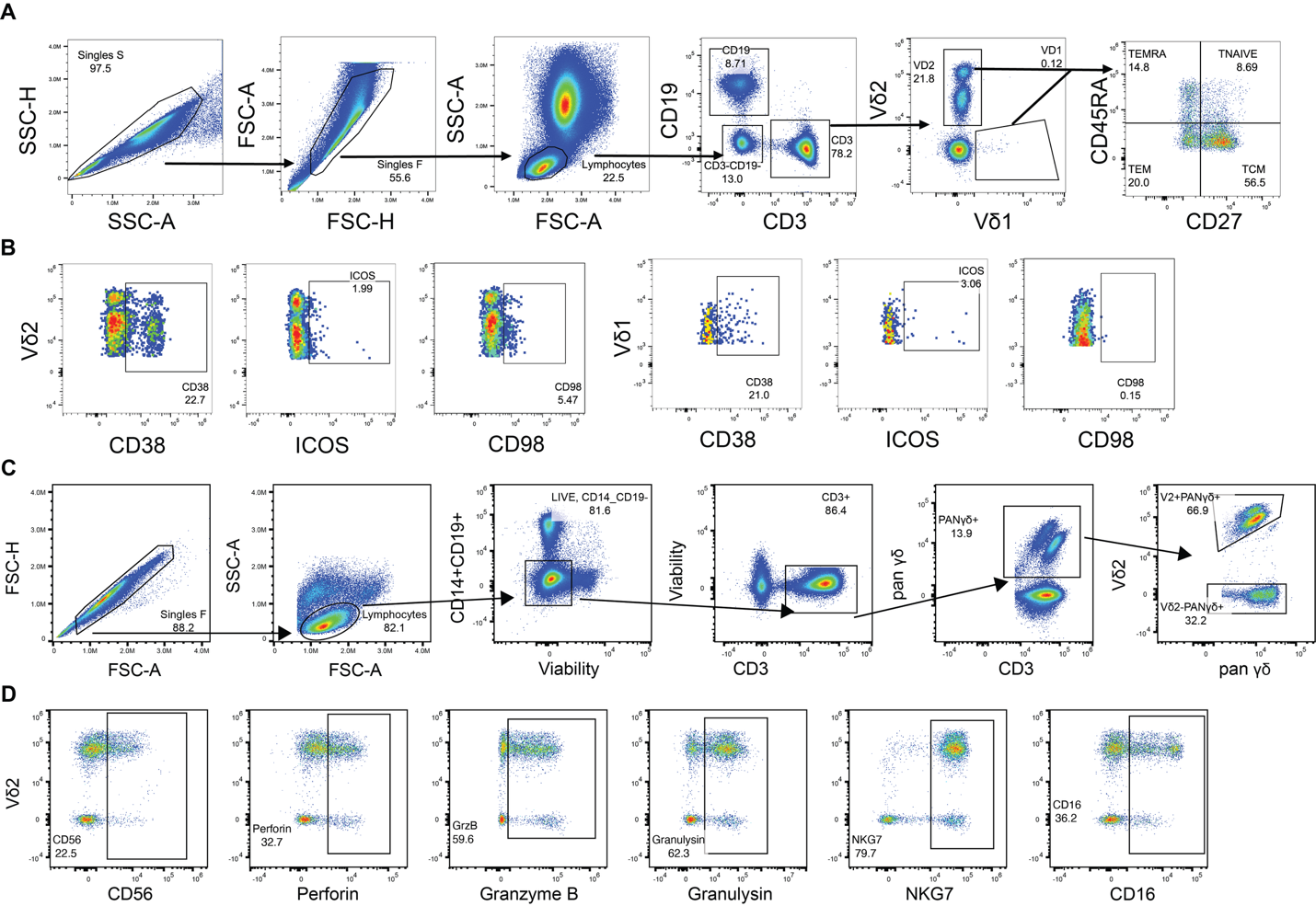

**Figure S3: *Ex vivo* analysis of γδ T cells during CHMI**

Gating strategies to investigated activation, proliferation and phenotypes of γδ T cells ex vivo in participants during CHMI with two flow cytometry panels. (**A**) Vδ2 and Vδ1 T cells were identified from CD3+ T cells (CD19-) live lymphocytes. Subsets were identified based on CD45RA and CD27 expression as naïve (CD45RA+CD27+), central memory (CM, CD45RA-CD27+), effector memory (EM CD45RA-CD27-) and TEMRA (CD45RA+CD27+). (**B**) Expression of activation markers CD38 and ICOS, and proliferation surrogate CD98 were quantified on each Vδ2 and Vδ1 T cells. (**C)** Phenotyping Panel 2: Vδ2+ and Vδ2- (largely consisting of Vδ1) cells were identified from pan γδ+CD3+ T cells (CD19-CD14-) live lymphocytes. (**D**) Expression of CD56, Perforin, Granzyme-B, Granulysin, NKG7, CD16 were quantified in γδ T cell subsets. Related to Figure 3.

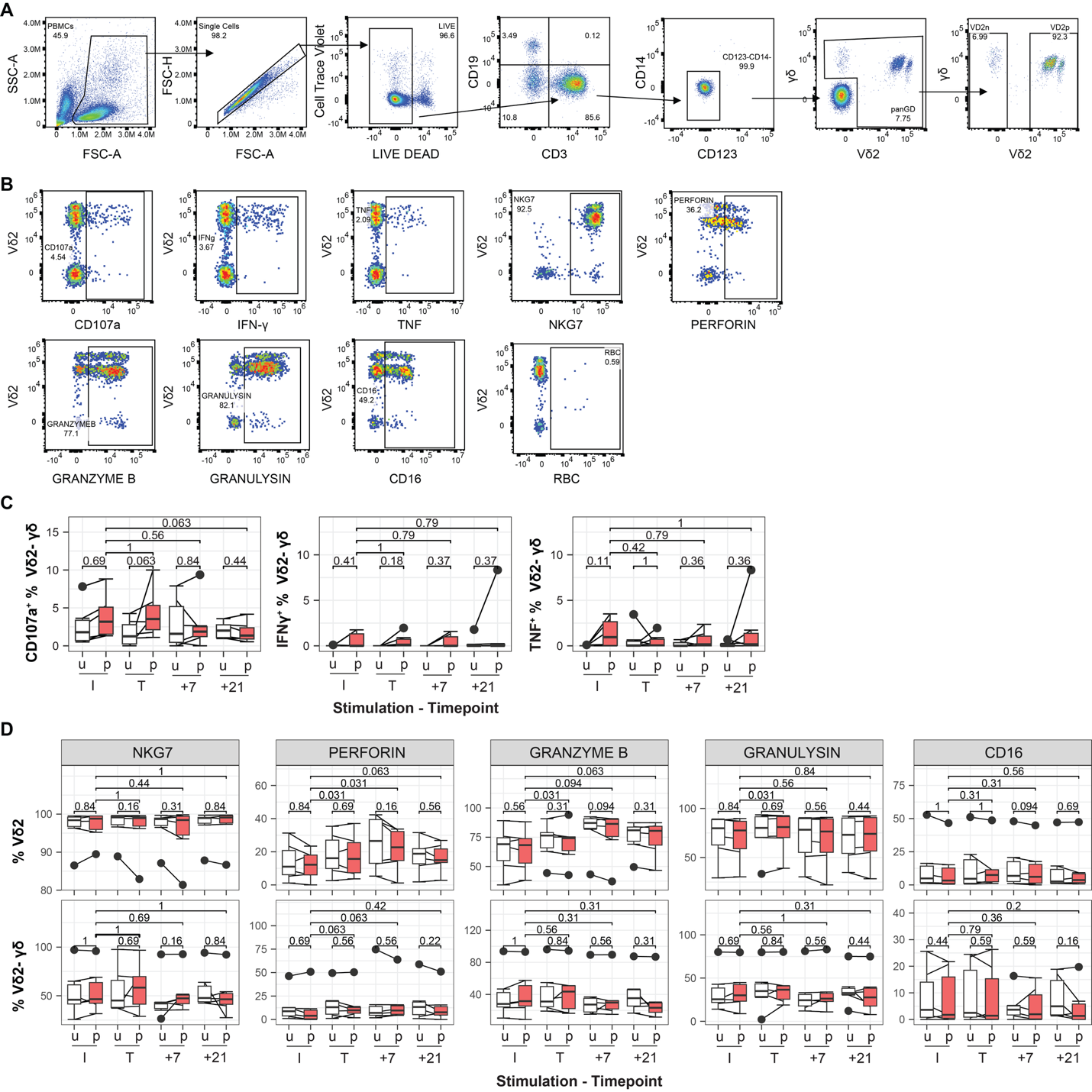

**Figure S4: *In vitro* analysis of γδ T cells during CHMI**

**(A/B)** Vδ2+ and Vδ2- γδ T were identified from γδ T cells (CD3+/CD19-/CD14-/CD123-/live lymphocytes), and functional capacity quantified by TNF, IFNγ expression, degranulation (CD107a), cytotoxic expression levels (NKG7, perforin, granzyme B, granulysin), CD16 expression and parasite uptake. **(C)** Vδ2- γδ T cell CD107a, IFNγ, TNF expression after stimulation with uninfected RBCs (u) or *Pf* infected RBCs (p) at infection (I, 0 days post infection (d.p.i.)), treatment (I, 8 d.p.i.) and at 15 d.p.i. (T+7) and 28 d.p.i. (T+20) in CHMI. (**D)** Expression of cytotoxic markers and CD16 in Vδ2+ and Vδ2- γδ T cells following stimulation with u or p across CHMI. Data are *n* = 6 individuals, box plots are show the median, box limits indicate the upper and lower quartiles, whiskers extend to 1.5 times the interquartile range. Lines represent paired observations. Comparisons made with Wilcoxon signed rank test. Related to Figure 3.

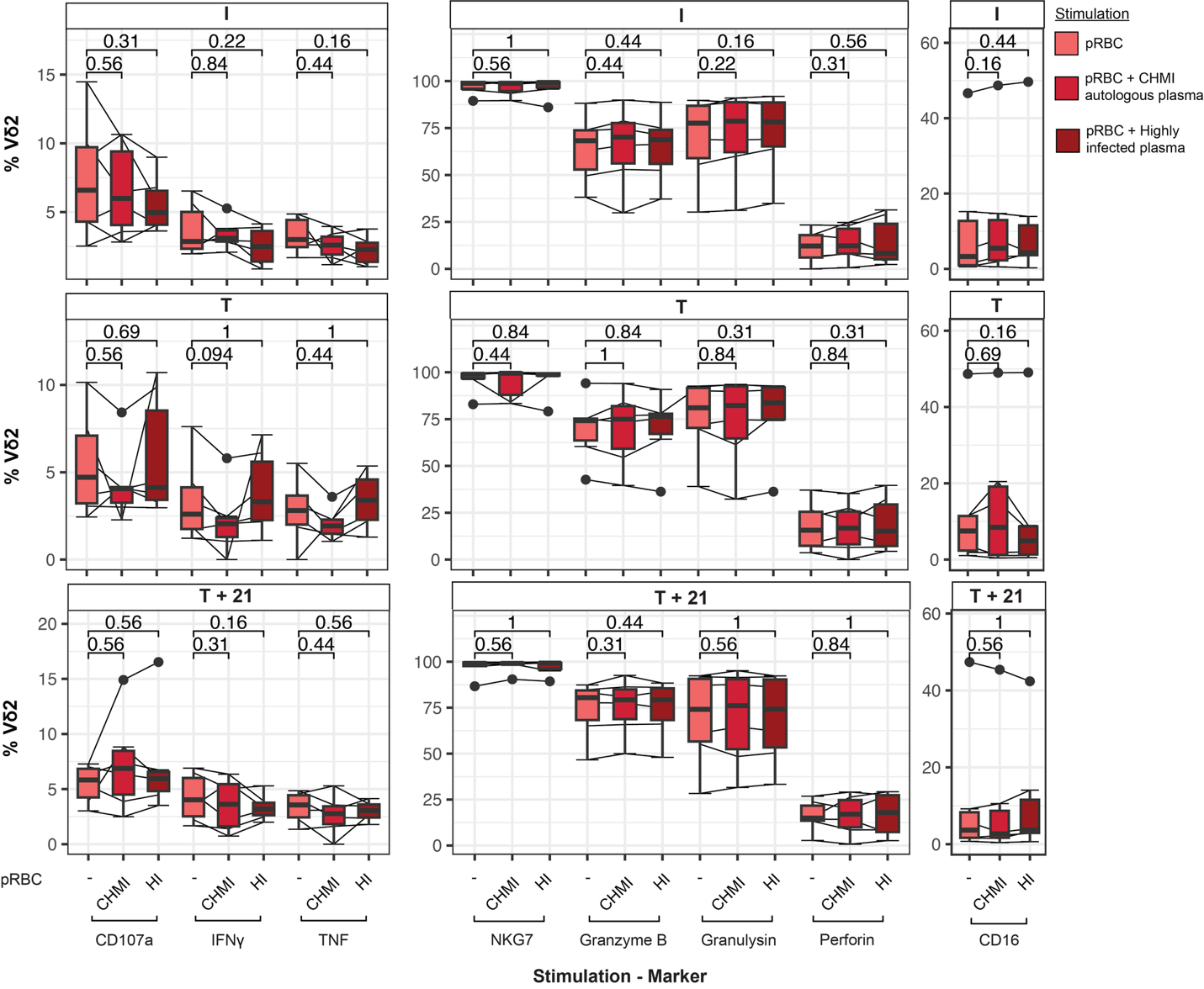
**Figure S5: *In vitro* antibody-dependent Vδ2 T cell responses in CHMI**

Parasite infected RBCs were unopsonised or opsonised with autologous plasma or plasma from highly exposed Ugandan donors and then incubated with PBMCs (n=6) from infection (I, 0 days post infection (d.p.i.)), treatment (I, 8 d.p.i.) and at 15 d.p.i. (T+7) and 28 d.p.i. (T+20) in CHMI. Expression of CD107a, IFNγ, TNF, NKG7, granzyme B, granulysin, perforin, CD16 and uptake of erythrocytes by Vδ2+ T cells from CHMI volunteers at day 0, 8 and 29. Box plots show the median, box limits indicate the upper and lower quartiles, whiskers extend to 1.5 times the interquartile range. Lines represent paired observations. Comparisons made with Wilcoxon signed rank test. See also Figure 3.

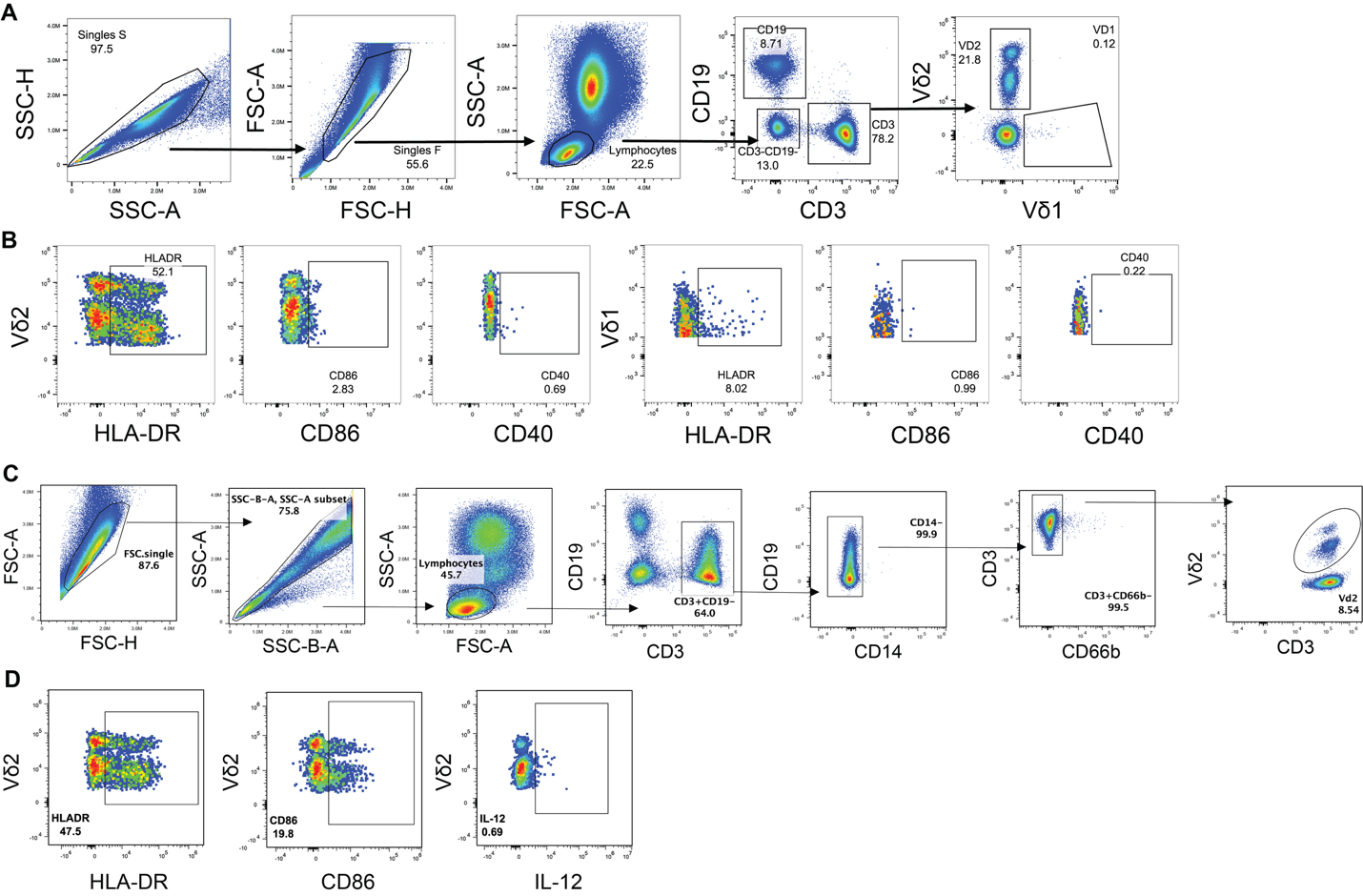

**Figure S6: *Ex vivo* analysis of APC-like γδ T cells during CHMI**

APC-like phenotypes of γδ T cells were examined *ex vivo* during CHMI. (**A**) Vδ2 and Vδ1 T cells were identified from CD3+ T cells (CD19-) single lymphocytes. (**B**) Expression of HLA-DR and co-stimulatory markers CD86 and CD40 were quantified on Vδ2 and Vδ1 T cells. Related to Figure 4.

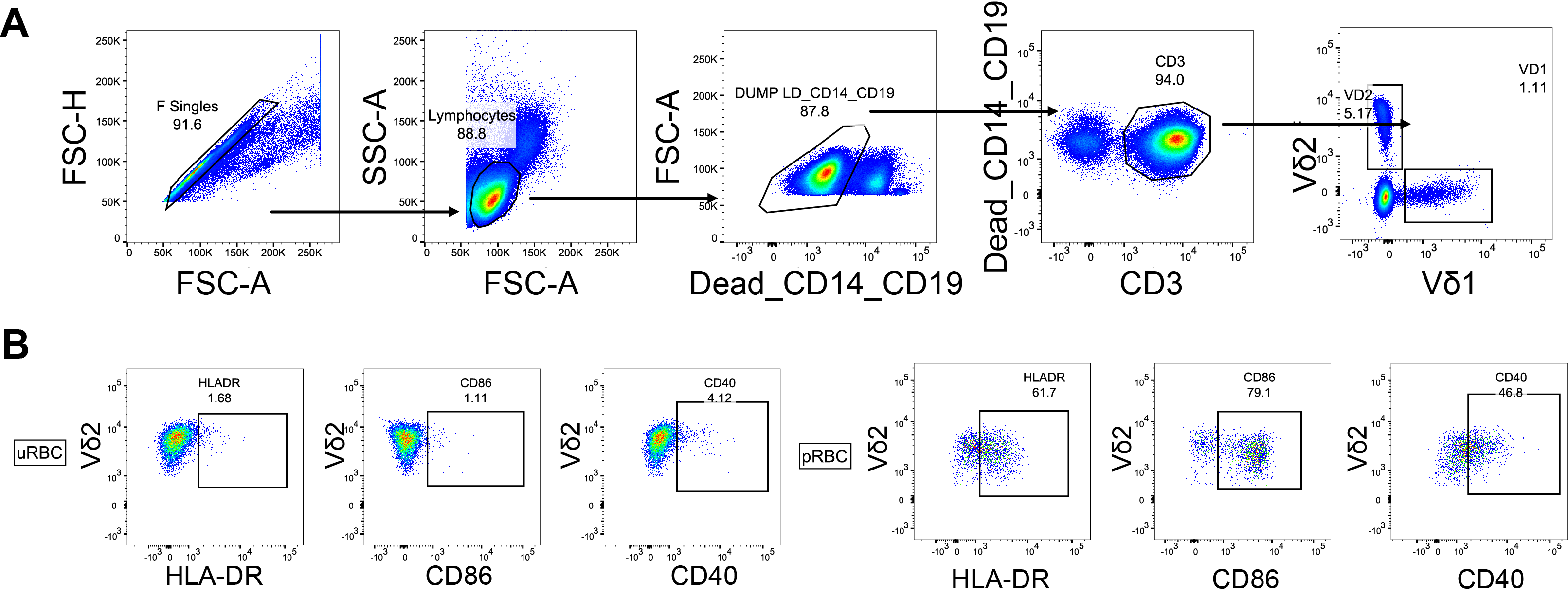

**Figure S7: Malaria-naïve γδ T cell APC-like phenotypes ex vivo and after co-culture**

Malaria-naive PBMCs (*n* = 15) were phenotyped ex vivo or after co-cultured with media alone, HMBPP, uRBC and pRBC for five days. (**A**) Representative gating strategy of Vδ2+ cells within the Single+/Lymphocyte+/ Dead-CD19-CD14-/CD3+ population. (**B**) Representative gating of HLA-DR, CD86 and CD40 in the Vδ2 T cell population after stimulation with uRBC or pRBC for five days. See also Figure 4.

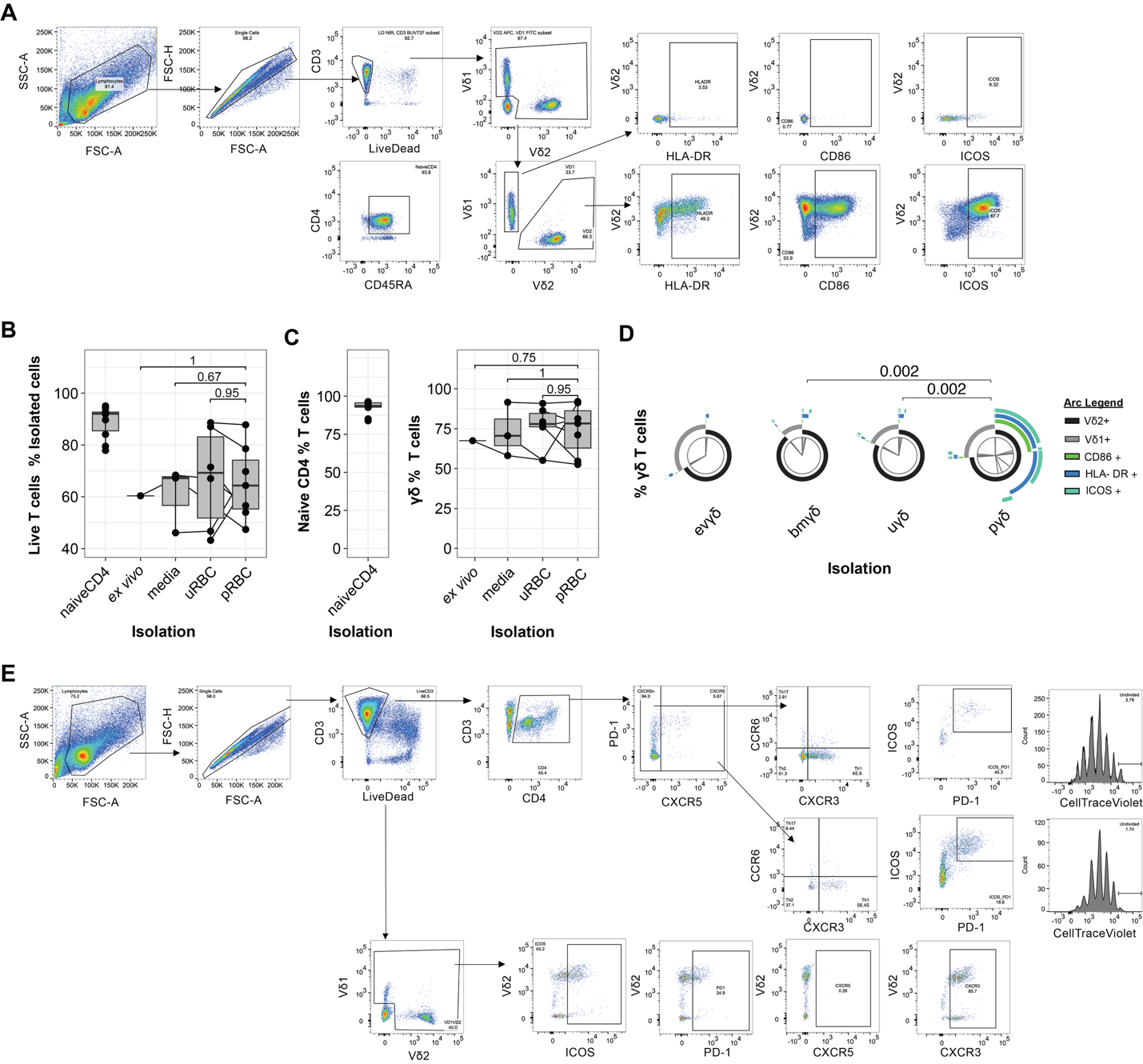

**Figure S8: γδ and CD4 T cells analysis for mixed lymphocyte reaction**

*Ex vivo* and *in vitro* cultured malaria-naive PBMCs were sorted with negative isolation bead kits to enrich naive CD4 T cells (*n* = 12) and γδ T cells (*n* = 6). *Ex vivo* naive CD4 T cells were co-cultured for six days with Human T-Activator CD3/CD28 Dynabeads^TM^ + recombinant cytokines IL-12, IL-23 and TGF-β (B+C), *ex vivo* isolated γδ T cells (*ex vivo*), γδ T cells pre-cultured with media alone (media), γδ T cells pre-cultured with uRBC (uRBC) or γδ T cells pre-cultured with pRBC (pRBC). (**A**) Representative gating of purified γδ and naïve CD4 T cells. (**B**) Viability of T cells after MACS isolation (**C**) Purity of CD4 and γδ T cells after MACS isolation. Data is median, box limits indicate the upper and lower quartiles, whiskers extend to 1.5 times the IQR. Lines are paired data. P is Wilcoxon signed rank test. (**D**) Induction of APC-like γδ T cells after culture and *ex vivo* in MACS isolations, shown by γδ T cell expression of Vδ2, Vδ1, CD86, HLA-DR and ICOS. Pie arc represent median frequencies and comparisons made by Permutation test. (**E**) Gating strategy do identify γδ and CD4 T cell phenotypes after mixed lymphocyte reaction. Related to Figure 4.

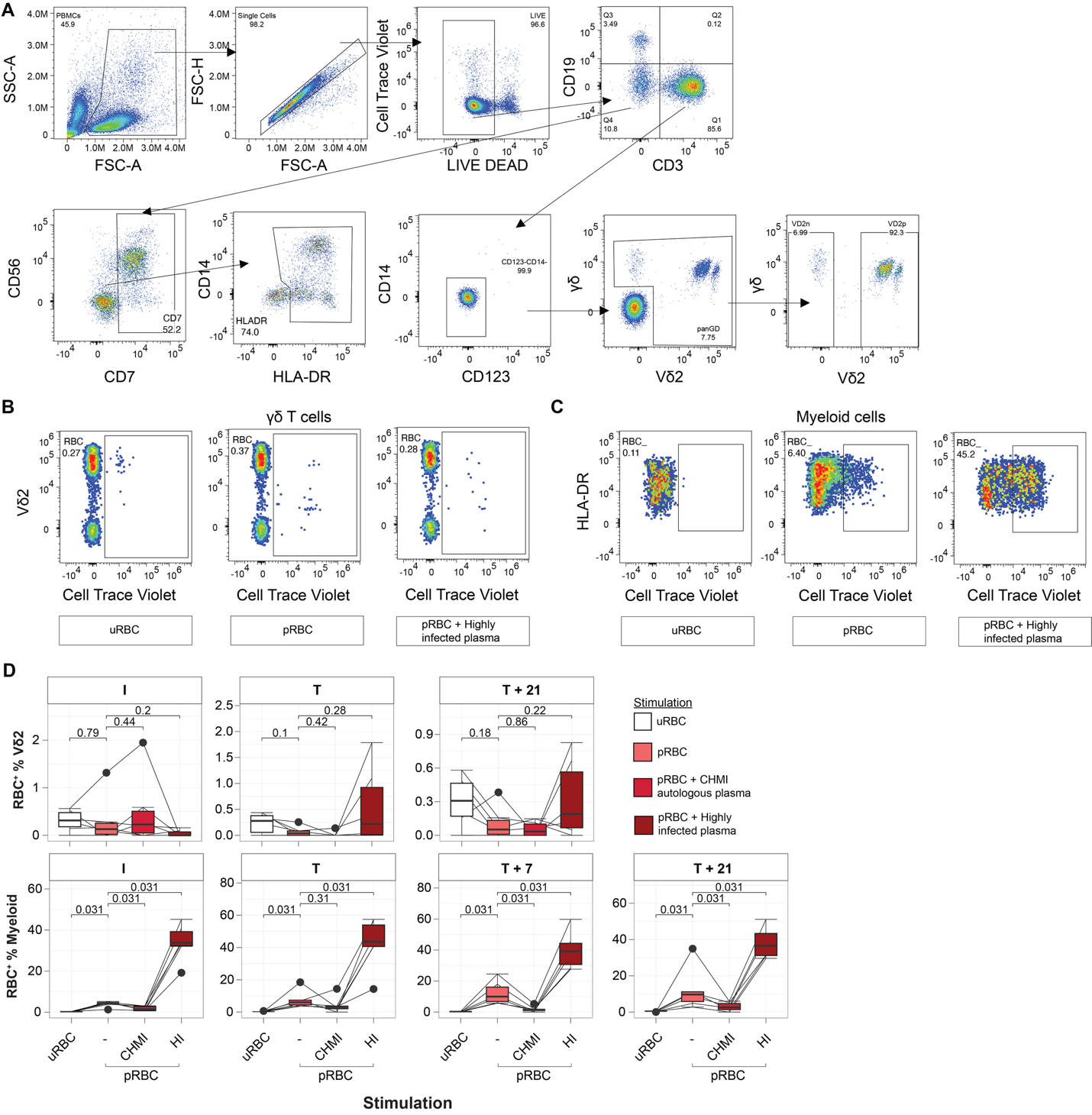

**Figure S9: Phagocytosis of RBCs by Vδ2 T and myeloid cells during CHMI**

Flow cytometric analysis of Vδ2 T and myeloid cells from CHMI participants following culture with Cell Trace Violet-stained uninfected (uRBC), and infected RBCs (pRBC) unopsonised or opsonised with autologous donor plasma (*Pf*-RBC + CHMI) and exposed (Ugandan-Highly Infected) donor plasma (pRBC + HI). (**A**) Gating strategy of γδ T (Lymphocyte+/Single+/Dead-/CD19-CD3+/CD14-CD123-/ γδ+/ Vδ2+) and myeloid (Lymphocyte+/Single+/Dead-/CD19-CD3-/HLA-DR+) cells. Representative Cell Trace Violet positive gates across conditions in (**B**) Vδ2 T and (**C**) myeloid cells. (**D**) Boxplots show frequency of Vδ2 T cells with internalized Cell Trace Violet of uRBC and pRBC. Centre lines represent median, box limits indicate the upper and lower quartiles, whiskers extend to 1.5 times the interquartile range. Lines represent paired observations. P is Wilcoxon signed rank test. Related to Figure 4.

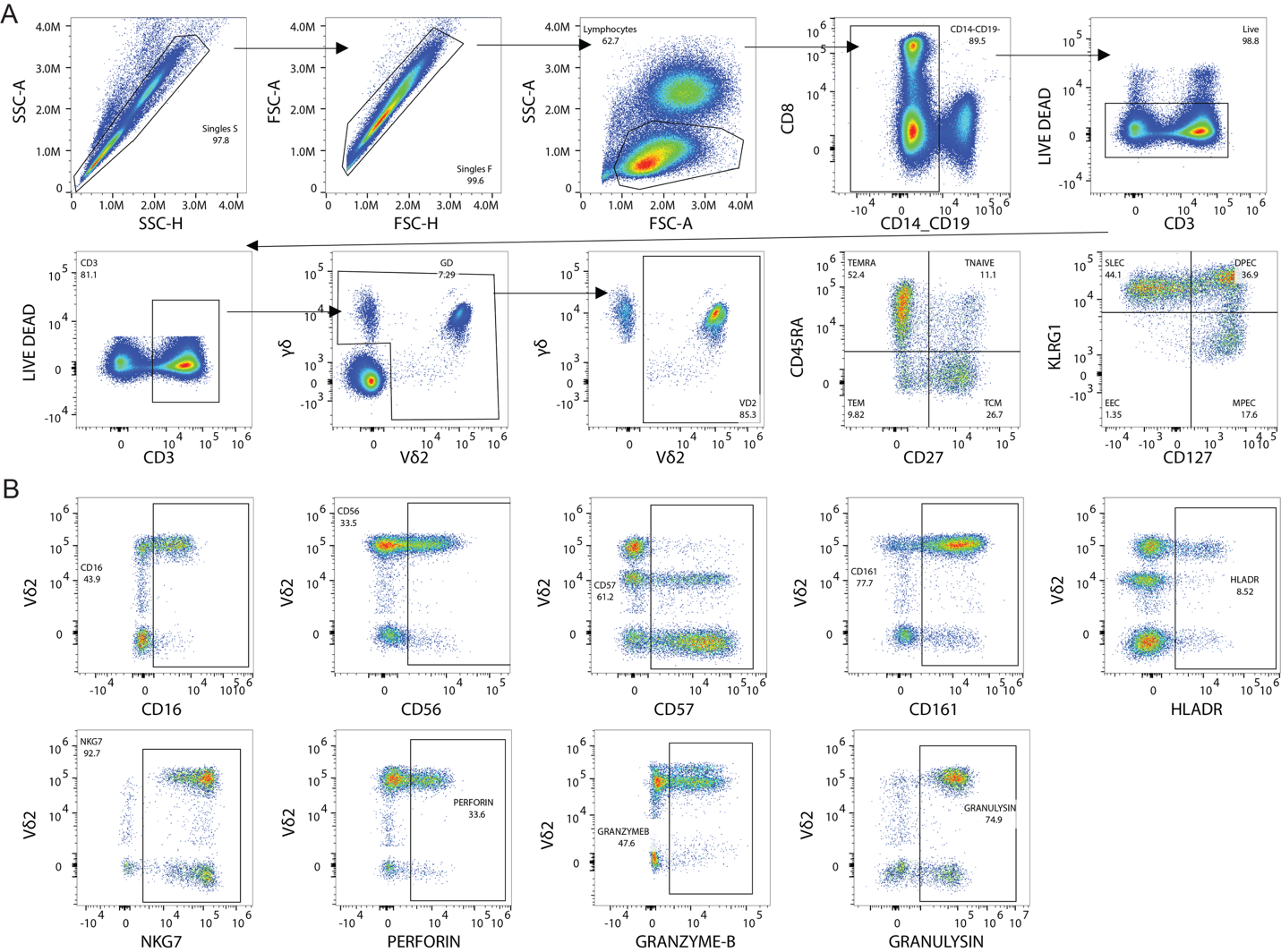

**Figure S10: Ruxolitinib CHMI cohort flow cytometric gating strategy**

(**A**) Vδ2+ and Vδ2- γδ T cells were identified from γδ+CD3+/CD19-/CD14-/live lymphocytes. γδ T cells memory subsets defined as TNAIVE (CD27^+^CD45RA^+^), TCM (CD27^+^CD45RA^-^), TEM (CD27^-^CD45RA^-^) and TEMRA (CD27^-^CD45RA^+^). γδ T cells effector subsets defined as DPEC [CD127^high^KLRG1^high^], MPEC [CD127^high^KLRG1^Low^], EEC [CD127^low^KLRG1^low^] and SLEC [CD127^low^KLRG1^high^]. (**B**) Positive gating example of CD16, CD56, CD57, CD161, HLA-DR, NKG7, perforin, granzyme B and granulysin by γδ T cells. Related to Figure 5.

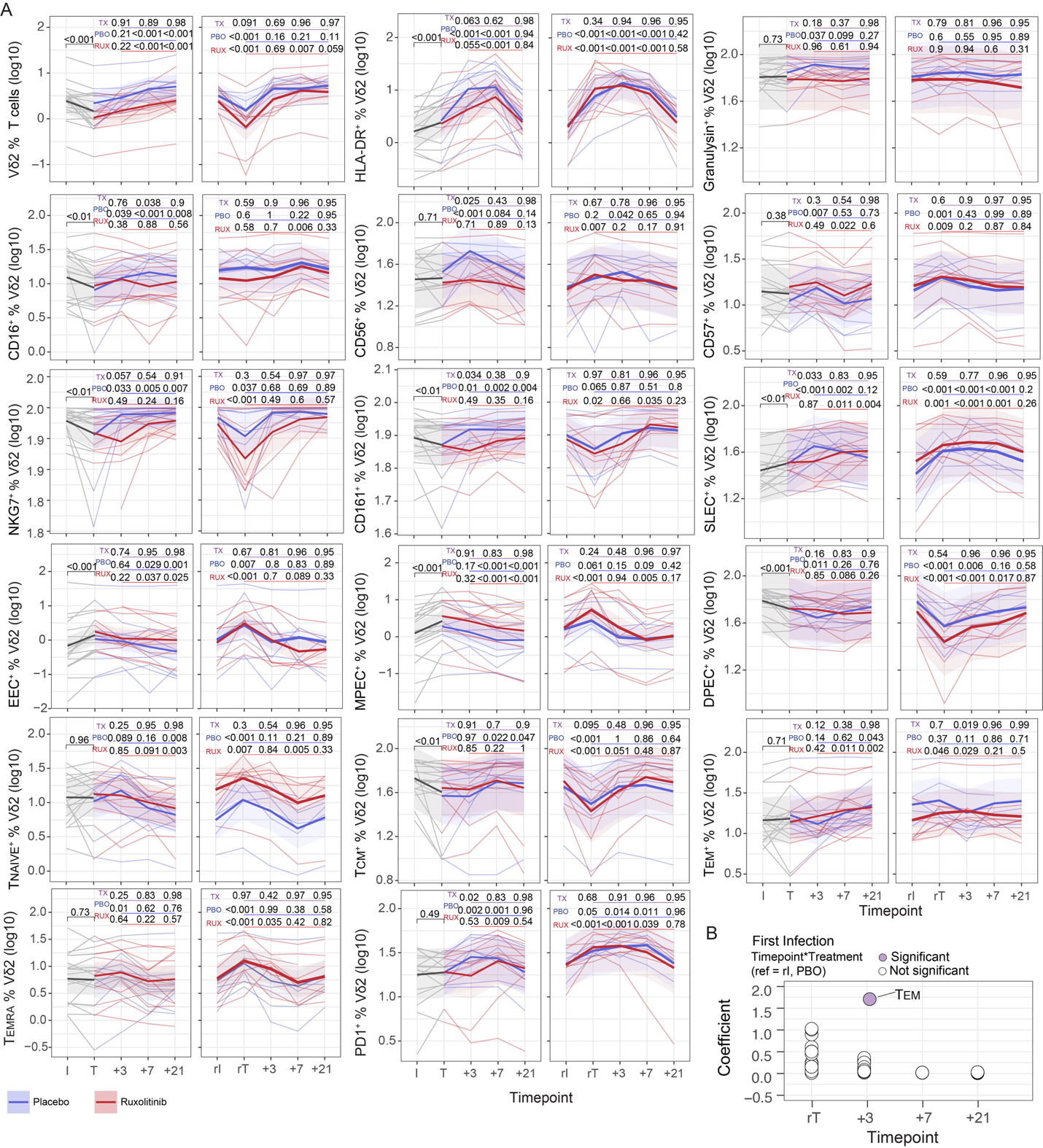
**Figure S11: Linear mixed model analysis of Vδ2^+^ γδ T cells in ruxolitinib CHMI**

(**A**) Marker expression by Vδ2^+^ γδ T cells during first and infection. Data are log-transformed with thin lines representing individuals and colored by treatment group (gray before inoculation, red in ruxolitinib-treated, and blue in placebo groups) and bold lines representing the mean of the predicted values from the fitted models for each group. P values (unadjusted) are from linear mixed-effect models. TX represents the P values for the interaction term between each time point (compared with time point T) and treatment group, i.e., the difference in change from baseline between the ruxolitinib and placebo groups (underlined in purple). P values for the comparison between each time point and time point T are shown for the placebo (PBO, underlined with blue) or ruxolitinib (RUX, underlined with red) group and were determined from contrasts. (**B**) Coefficient of TX from linear models of all Vδ2^+^ T parameters in second infection. Related to Figure 5.

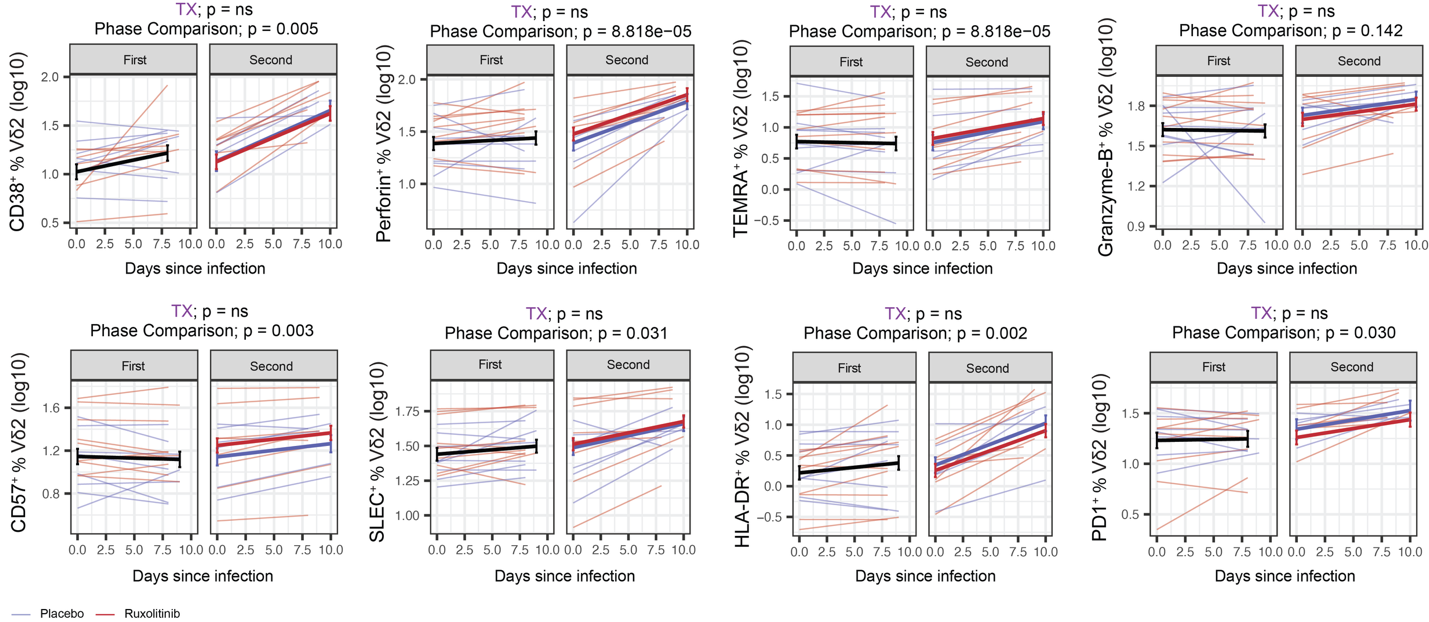
**Figure S12: Kinetics of Vδ2^+^ γδ T cell activation during first and second infection.**

(**A**) Vδ2^+^ γδ T cells activation in first and second infection (expression of CD38, Perforin, Granzyme-B, CD57, HLA-DR and PD1 and proportions of SLEC and TEMRA) analysed with log-linear mixed effects models to identify changes in activation between from I to T in first and rI and rT in second CHMI. TX is the interaction [all nonsignificant] term for an impact of treatment in first infection on the change in induction of response between infections. Phase Comparison is the P value of the change in slope between first and second infection. Related to Figure 5.

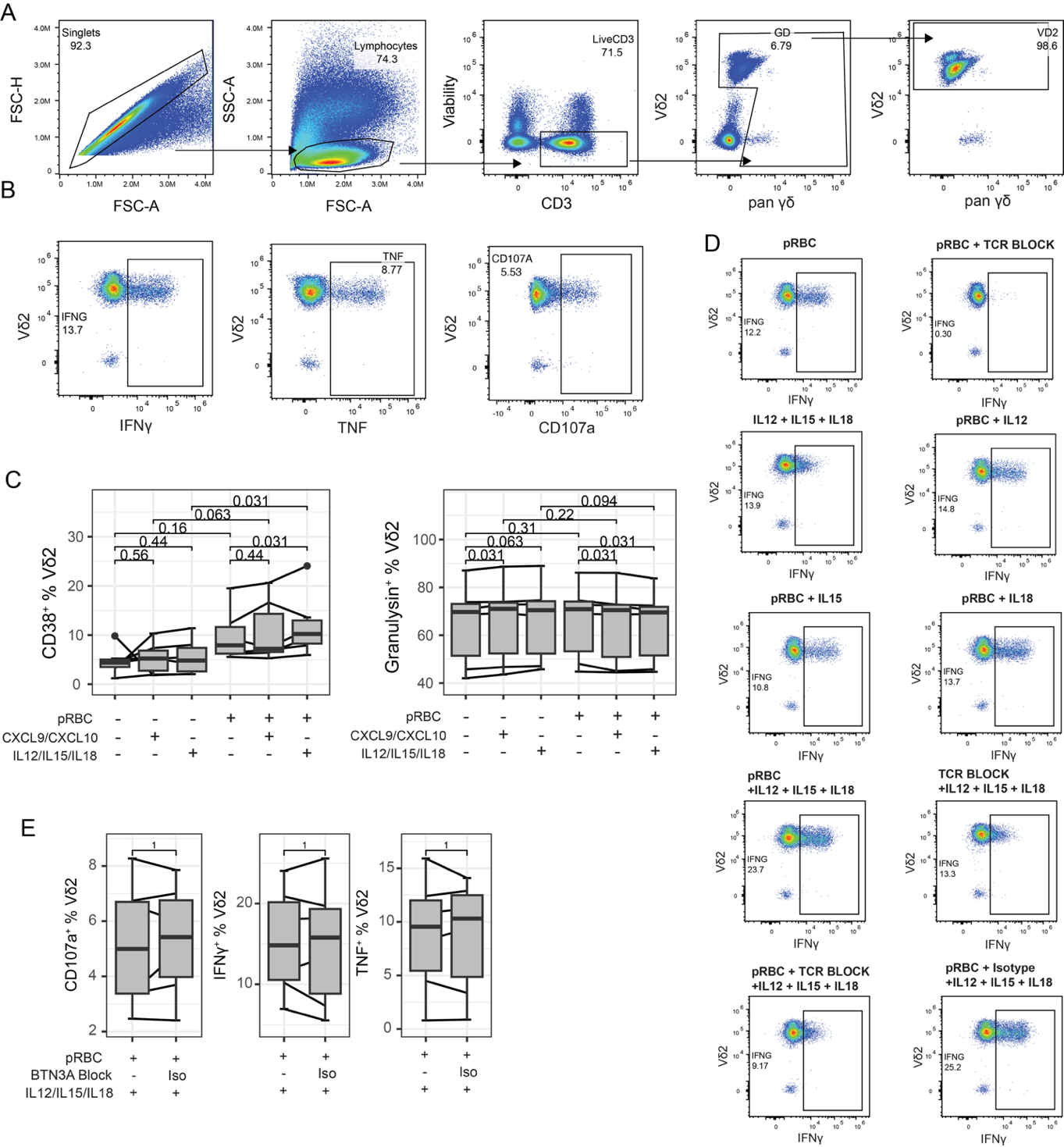

**Figure S13: Cytokine and TCR activation of Vδ2 T cells in vitro**

PBMCs were isolated from malaria-naïve donors (n=6) and co-cultured with *Pf*-lysate or media with and without CXCL9/10 or IL-12/15/18, blocked with mAb 103.2 (BTN3A Block) or an isotype control. Gating strategy to identify Vδ2+ cells and CD107a, IFNγ and TNF expression. (**C**) CD38 and Granulysin expression by Vδ2 T cells after co-culture with *Pf*-lysate or media with and without CXCL9/10 or IL-12/15/18. (**D**) Representative plots of CD107a, IFNγ and TNF on γδ T cells in each condition. (**E**) CD107a, IFNγ and TNF expression by Vδ2 T cells after co-culture with *Pf*-lysate and IL-12/15/18 with no mAb 103.2 (BTN3A Block) or an isotype control. Centre lines represent median, box limits indicate the upper and lower quartiles, whiskers extend to 1.5 times the interquartile range. Lines represent paired observations. Paired comparisons made with Wilcoxon signed rank test. Related to Figure 6.

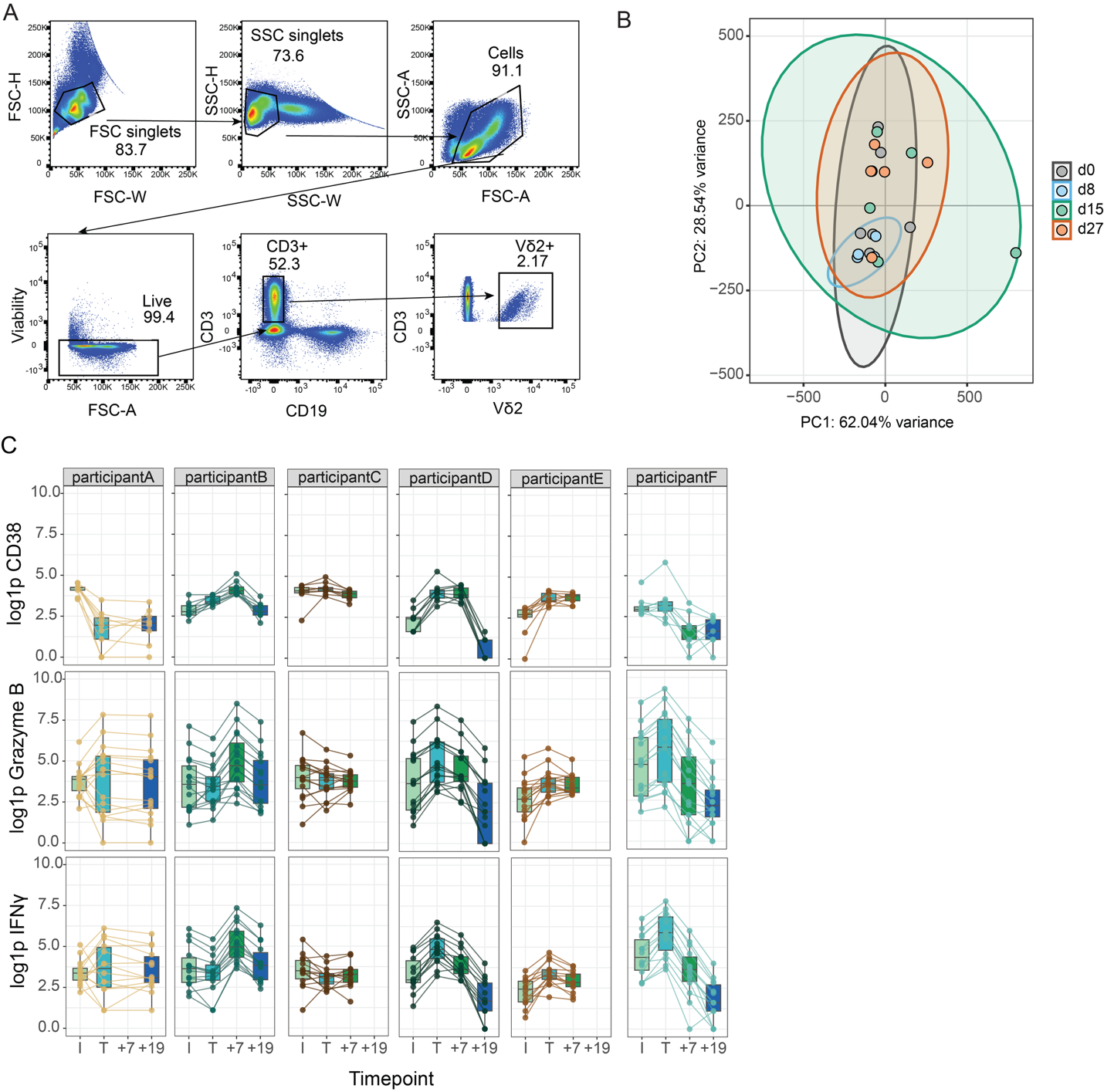
**Figure S14: Vδ2 T cells ATAC-seq analysis**

(**A**) Vδ2 T cells flow gating strategy to sort Single+/Cells+/Dead-/CD3+CD19-/Vδ2+ cells for ATAC-seq. (**B**) Principal Component Analysis of peak counts PC1 v PC2 by timepoint from all participants. (**C**) Log1p Peak Counts of CD38, granzyme B and IFNγ gene regions in participants across timepoints.

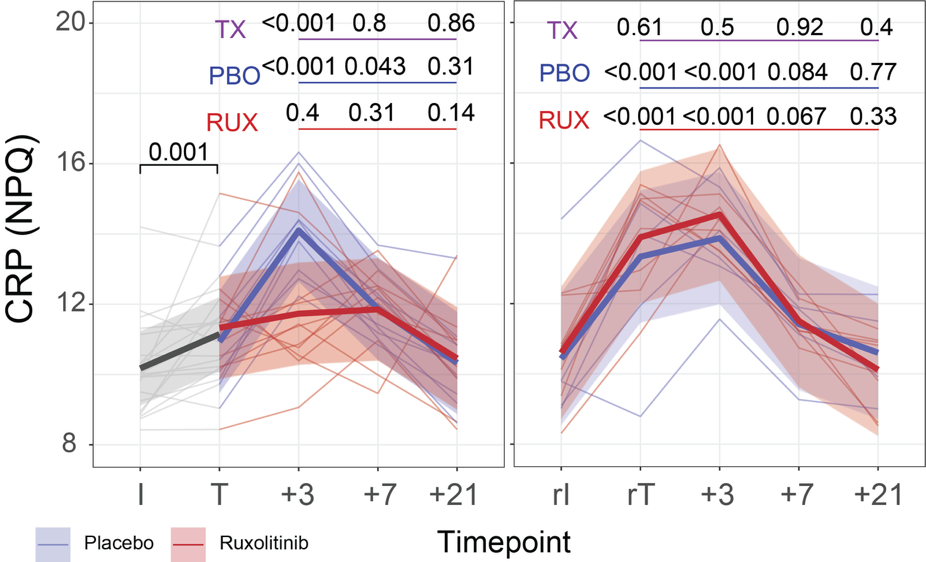

**Figure S15: C-reactive protein levels in Ruxolitinib CHMI**

C-reactive protein (CRP) levels (Nulisa Protein Quantification) in volunteers (n=20) during CHMI to test ruxolitinib as a host-directed therapy. Individuals were infected (I), and at treatment with anti-malaria drugs and ruxolitinib or placebo at (T, 8/9 d.p.i.), and then followed (T+3, T+7, T+21). At day 91 individuals were reinfected and cells analysed (rI, rT, rT+3, rT+7, rT+21). Data are from NUlisa-seq with thin lines representing individuals and colored by treatment group (grey before inoculation, red in ruxolitinib-treated, and blue in placebo groups) and bold lines representing the mean of the predicted values from the fitted models for each group. P values (unadjusted) are from linear mixed-effect models. TX represents the P values for the interaction term between each time point (compared with time point T) and treatment group, i.e., the difference in change from baseline between the ruxolitinib and placebo groups (underlined in purple). P values for the comparison between each time point and time point T are shown for the placebo (PBO, underlined with blue) or ruxolitinib (RUX, underlined with red) group and were determined from contrasts. Related to Figure 7.
